# Smoking and SARS-CoV-2 Impair Dendritic Cells and Regulate DC-SIGN Expression in Tissues

**DOI:** 10.1101/2020.12.23.20245316

**Authors:** Guoshuai Cai, Yohan Bossé, Mulong Du, Helmut Albrecht, Fei Qin, Xuanxuan Yu, Xizhi Luo, Michelle Androulakis, Xia Zhu, Jun Zhou, Xiang Cui, Changhua Yi, Chao Cheng, Mitzi Nagarkatti, Prakash Nagarkatti, David Christiani, Michael Whitfield, Christopher Amos, Feifei Xiao

## Abstract

The current spreading novel coronavirus SARS-CoV-2 is highly infectious and pathogenic. In this study, we screened the gene expression of three SARS-CoV-2 host receptors (ACE2, DC-SIGN and L-SIGN) and DC status in bulk and single cell transcriptomic datasets of upper airway, lung or blood of smokers, non-smokers and COVID-19 patients. We found smoking increased DC-SIGN gene expression and inhibited DC maturation and its ability of T cell stimulation. In COVID-19, DC-SIGN gene expression was interestingly decreased in lung DCs but increased in blood DCs. Strikingly, DCs shifted from cDCs to pDCs in COVID-19, but the shift was trapped in an immature stage (CD22+ or ANXA1+ DC) with MHCII downregulation in severe cases. This observation indicates that DCs in severe cases stimulate innate immune responses but fail to specifically recognize SARS-CoV-2. Our study provides insights into smoking effect on COVID-19 risk and the profound modulation of DC function in severe COVID-19.

**Graphical Abstract:** 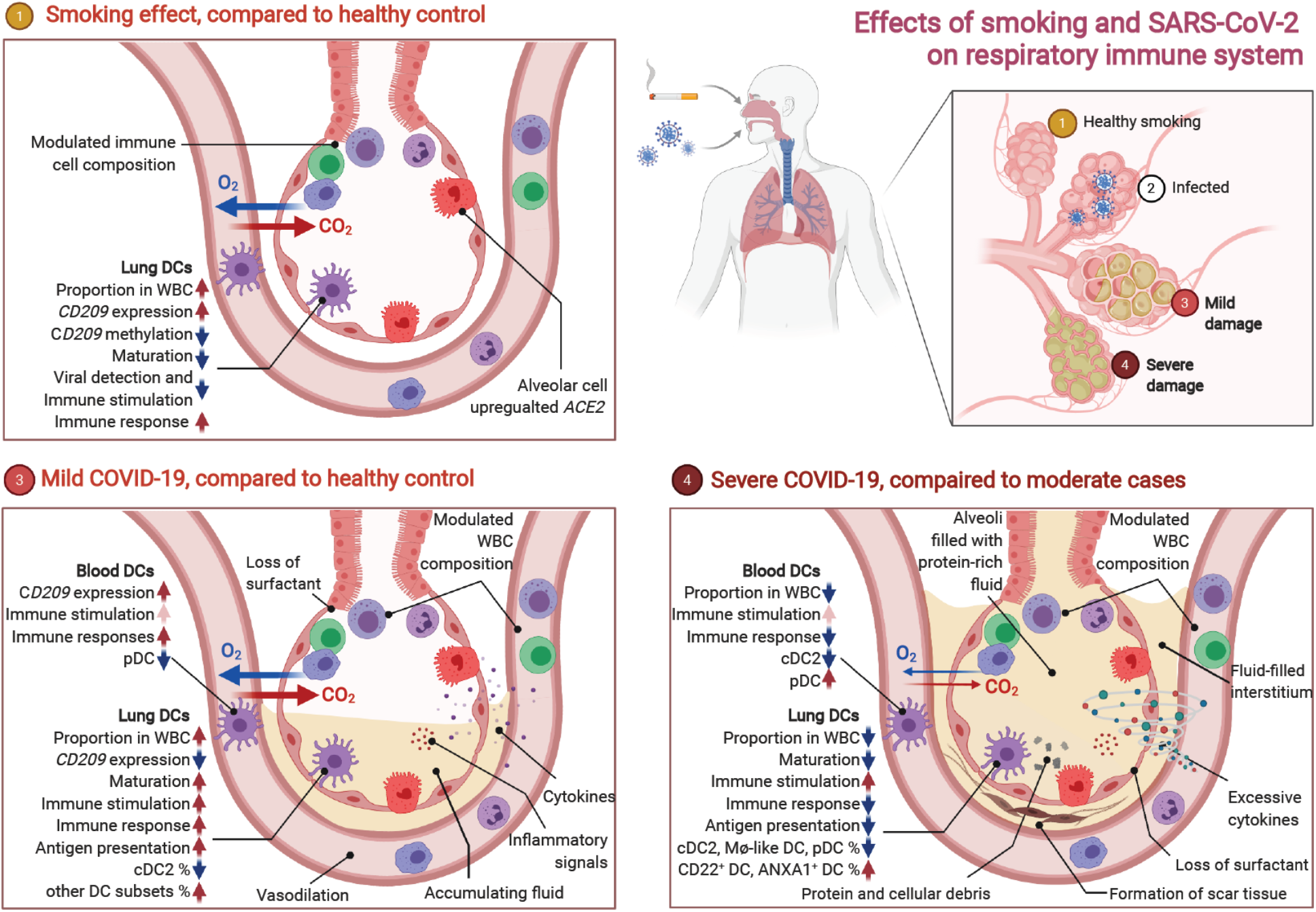

**Highlights:** Smoking upregulates the expression of *ACE2* and *CD209* and inhibits DC maturation in lungs. SARS-CoV-2 modulates the DCs proportion and *CD209* expression differently in lung and blood.

Severe infection is characterized by DCs less capable of maturation, antigen presentation and MHCII expression.

DCs shift from cDCs to pDCs with SARS-CoV-2 infection but are trapped in an immature stage in severe cases.

## Introduction

In 2020, a novel coronavirus SARS-CoV-2 has been spreading as a pandemic infection. SARS-CoV-2 is highly infectious and pathogenic through human-to-human transmission and causes severe Coronavirus Disease 2019 (COVID-19)^1^. COVID-19 critical cases are often characterized by a pro-inflammatory “cytokine storm” with a release of excessive cytokines (including IL-6, IL-1, IL-2, IL-10, TNF-*α* and IFN-*γ* and others) and a hyperactive immune response which cause damage to target organs. Despite extensive research, the exact immune pathophysiology of SARS-CoV-2 has remained elusive.

Coronavirus is a single-stranded RNA virus that can be divided into four main genres including the alpha, beta, gamma and delta coronaviruses^2^. Studies showed that SARS-CoV-2 is closely related to SARS-CoV, with around 80% homology of genome^3^, belonging to the same *β*-genus^4^, having similar receptor-binding domain (RBD) structures^2^ and causing similar clinical symptoms including acute respiratory response^5^. Many viruses are capable of using alternative receptors to enter host cells. This is also true for SARS coronavirus. Three receptors—ACE2, DC-SIGN and L-SIGN (gene symbol as *ACE2, CD209* and *CLEC4M* respectively) have been found to be involved in the pathogenicity of SARS-CoV^6-8^. ACE2 was also been quickly confirmed to be the receptor for SARS-CoV-2^9^. Recently, DC-SIGN and L-SIGN were identified as alternative receptors for mediating SARS-CoV-2 entry into human cells^10^.

In vivo, L-SIGN is largely expressed on endothelial cells in liver sinusoids and lymph nodes, whereas DC-SIGN is mainly expressed on dendritic cells (DCs)^11^. Aside from epithelial cells, SARS-CoV infection was found in T cells, macrophages (Mø) and monocyte-derived dendritic cells and this viral attack is probably responsible for the lymphopenia which was commonly observed in patients with SARS-CoV infection^12^. While DC-SIGN and L-SIGN provide gateways for SARS coronavirus to attack infected immune cells, they play important anti-viral roles. They both capture, transmit and disseminate virus within the host^13,14^. With the dual roles of DC-SIGN as a SARS-CoV-2 gateway and an innate immune initiator, DCs may play an important and complicated role in OVID-19 patients and link to clinical phenotypes. Studies indicate a decrease of conventional DCs (cDC) and plasmacytoid DCs (pDC) in the lung and blood of severe COVID-19 cases^15,16^. Yet, comprehensive insights into modulation of DCs in tissues with COVID-19 are still missing.

Environmental exposures might influence the COVID-19 risk by regulating receptors and remodel immune cells. The clinical epidemiology data showing a higher risk of smokers of developing a severe COVID-19 disease^17^. In agreement with that, we previously demonstrated an upregulation of ACE2 in the lungs of smokers^18^. In this study, we further investigated the effects of smoking on the gene expression of the two newly identified SARS-CoV-2 receptors DC-SIGN and L-SIGN by analyzing large-scale bulk tissue datasets and high-granularity single cell datasets of normal lung tissues. Also, we systematically studied the transcriptome dysregulation in single cells in the upper airway (nasopharynx/pharynx samples), lower airway (bronchoalveolar lavage fluid (BALF) samples) and peripheral blood (peripheral blood mononuclear cell (PMBC) samples) of COVID-19 patients. These single cell datasets enabled the investigation of variation of DC activation in smokers and non-smokers, as well as mild and severe COVID-19 and healthy controls. In severe COVID-19, we found significantly decreased DCs and its shift to immature subsets. The results illustrate the pathogenicity and virulence of SARS-CoV-2 infection from the aspect of DCs which is a critical player in the immune response to viral infection.

## Results

### Infection correlation and variation of immune factors in COVID-19 upper airway

In the nasopharynx/pharynx samples of 238 patients with acute respiratory illnesses (ARIs), we found ACE2 (P = 3.9 × 10^−10^) and DC-SIGN (*P* = 0.048) gene expression were significantly higher with the SARS-CoV-2 infection (Fig. S1). In addition, we found *ACE2* gene expression was strongly positively associated with the viral load of SARS-CoV-2 (*P* = 5.3 × 10^−4^). No data was available for *CLEC4M*. Notably, the gene expression of essential factors of antigen-presenting cells (APC) for antigen presentation including CD40, CD80 and CD86, HLA genes, IgG immunoglobulin receptor (FcR) III and interferon-gamma (IFN-*γ*) were upregulated in SARS-CoV-2 ARIs compared to non-viral ARIs, whereas IgA and IgE FcRs, interferon-kappa (IFN-*κ*), and cyclooxygenase (COX) genes *PTGS1* and *ALOS5* were downregulated. The gene expression of ACE2, CD80, CD86, CD83, IFN-*γ*, FCER1G and IgG FcRs were increasingly associated with the viral load of SARS-CoV-2 (Fig. 1A, Fig. S1). No such association were observed in HLA genes. In addition, the proportions of activated CD4^+^ memory T cells (CD4m T) and M1 Mø were increased in SARS-CoV-2 ARIs and significantly associated with increased viral load (Fig. 1A, Fig. S1), while the proportions of activated mast cells and neutrophils were decreased. The proportions of these 4 cell types and the active DC well predicted the SARS-CoV-2 load (*ρ* = 0.79, *P* = 1.3 × 10^−7^, Fig. 1A) and discriminated between non-viral and SARS-CoV-2 ARIs (*P* = 1.2 × 10^−5^, Fig. S1).

**Figure 1.**
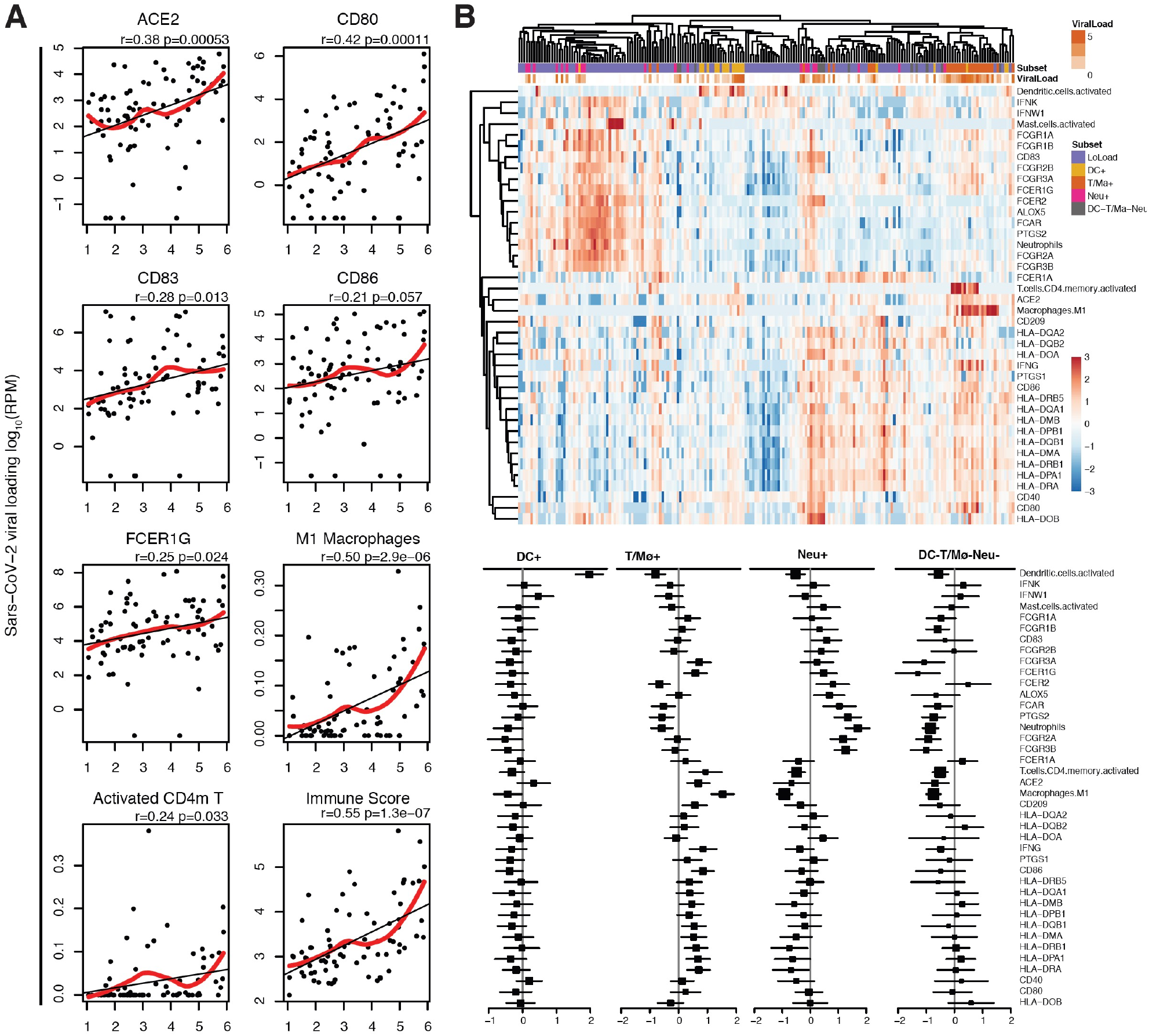
Correlation and variation of SARS-CoV-2 receptors and immune factors in nasopharynx/pharynx samples of SARS-CoV-2 ARIs. (A) Scatter plots of gene expression profiles of SARS-CoV-2 receptor ACE2, immune response activators (CD80, CD83 and CD86), the proportion of active CD4 memory T cells and M1 macrophages in 22 immune cell types, as well as scores from a linear model of 5 immune cell types (active CD4 memory T cells, M1 Mø, mast cells, neutrophils and active DCs). Their associations with viral load are also visualized in scatter plots. The black and red lines are fits of linear model and LOESS, respectively. (B) Hierarchical clustering and heatmap of SARS-CoV-2 receptors, immune factors and proportions of 5 immune cell types in ARI subsets, including 1 subset with low SARS-CoV-2 load (LoLoad) and 4 subsets with high SARS-CoV-2 load and activated DCs (DC+), activated T or M1 Mø (T/Mø+), activated neutrophiles (Neu+) and DC-T/Mø-Neu-. Compared each subset to others, the fold changes and 95% confidence intervals of each feature are shown on the bottom. The size of the squares is proportional to the standard deviation.

Interestingly in ARIs with a significant SARS-CoV-2 viral load (log_10_ CPM > 1), we observed distinct subtypes associated with activation of DCs and T/Mø cells (Fig. 1B). According to the activation of specific immune cell types, we further classified ARIs into four subtypes DC+ and T/Mø+, Neu+ and DC-T/Mø-Neu-(Fig. 1B). The DC+ subtype was characterized with activated DCs, downregulated FcRs and HLA genes. The T/Mø+ subtype was characterized with activated T or Mø, upregulated viral gateway *ACE2* and *CD209* as well as immune factors including *INFG, CD86, FCGR3A, FCGR1G and* HLA genes. The Neu+ subtype was characterized with enriched neutrophils, upregulated FcRs genes and Cox genes (*PTGS2* and *ALOS5*), and downregulated HLA genes and *ACE2*. The DC-T/Mø-Neu-subtype was characterized with downregulated FcRs genes, *ACE2* and *CD209*. These results provide new clues for the variation of immune response in COVID patients with its potential link to the activation of specific immune cell types. Not like the T/Mø+ subtype with activated immune factors and high expression of *ACE2* and *CD209*, DC+, Neu+ and DC-T/Mø-Neu-subtypes may produce high virus load by immunodeficiency.

### DC-SIGN is associated with COVID-19 severity, immune- and neural-related phenotypes and respiratory diseases

We investigated the association of the expression of ACE2, DC-SIGN and L-SIGN with COVID-19 risk by large-scale population genetic association analyses (see Methods). Using Mendelian Randomization (MR), we found that the expression of DC-SIGN plasma protein were associated with increased COVID-19 risk (*β* = −0.14, *P* = 3.39 × 10^−3^) and severity (*β* = −0.30, *P* = 5.41 × 10^−5^; Table S1). Such significant associations were not found in ACE2 and L-SIGN.

The important role of DC-SIGN in phagocytosis and immune activation was also reflected by the phenotype associations detected in this study. GWAS found that the genetic variation of *CD209* is associated with multiple diseases, including asthma (the top among detected associations, *P* = 1.0 × 10^−13^), cancers and neurological disorders (Table S2), showing the possible underlying links involving by DCs and/or Mø. Neurological symptomatology associated with COVID-19 includes headaches, confusion, hyposmia, encephalitis, myelitis, neuropathy, and stroke^19,20^. Given circulating DCs in cerebrospinal fluid are capable of reaching cervical lymph nodes outside central nervous system (CNS)^21^, our data suggests DCs is a possible route for SARS-CoV-2 infection in the brain, which also provides a new explanation to the neurotropism of SARS-CoV-2^22^. Moreover, the transcriptome-based PheWAS analysis found that *CD209* gene expression is significantly associated with the immune cell composition, including the depletion of neutrophils (count: *β* = −0.10, *P* = 4.0 × 10^−5^; percentage: *β* = −0.60, *P* = 3.2 × 10^−5^) and overall WBC (count: *z* = −3.34, *P* = 8.3 × 10^−4^), as well as the augmentation of lymphocytes (percentage: *β* = 0.14, *P* = 3.00 × 10^−4^) and monocytes (percentage: *β* = 0.15, *P* = 1.24 × 10^−3^) in lung (Table S3). These associations were further confirmed by MR analysis on DC-SIGN plasma protein expression (neutrophil count: *β* = −0.03, *P* = 1.38 × 10^−16^; neutrophil percentage: *β* = −0.02, *P* = 0.02; WBC count: *β* = −0.03, *P* = 5.40 × 10^−27^; lymphocyte percentage: *β* = 0.02, *P* = 5.29 × 10^−5^; monocyte percentage: *β* = 0.03, *P* = 8.52 × 10^−3^; Table S4). Moreover, we also observed such significant associations of *CD209* expression with immune cell composition in blood samples of 160 ARDS cases and 142 non-ARDS controls, regardless of the disease status (Fig. S2). In addition, we found that *CD209* expression was associated with immune related symptoms such as tiredness or low energy, depression and duration of fitness (Table S3). In lung adenocarcinoma tumors, we only observed a slight downregulation of *CD209* (*beta* = −0.26, *P* = 0.07) and no significant association with survival (*P* = 0.33) and tumor stage (*P* = 0.80).

### Smoking upregulates the expression and decreases the methylation of CD209 in lungs

We previously have found that smoking upregulates the lung expression of *ACE2*^18^. In this study, we further investigated the effect of smoking on expression of *CD209* and *CLEC4M* from 8 independent studies (N=1162). Adjusted for age and sex, *CD209* expression was significantly upregulated in smoker lungs (meta-analysis *β* = 0.15, *P* = 1.1 × 10^−4^, Fig. 2A). Significant association (meta-analysis *β* = 0.1, *P* = 0.03) between *CD209* expression and smoking status (never-, former- and current-smoker) was also observed with 4 out of 5 available datasets showing a positive trend of association (Fig. 2A). We did not find a significant difference in *CLEC4M* gene expression (Fig. S3).

**Figure 2.**
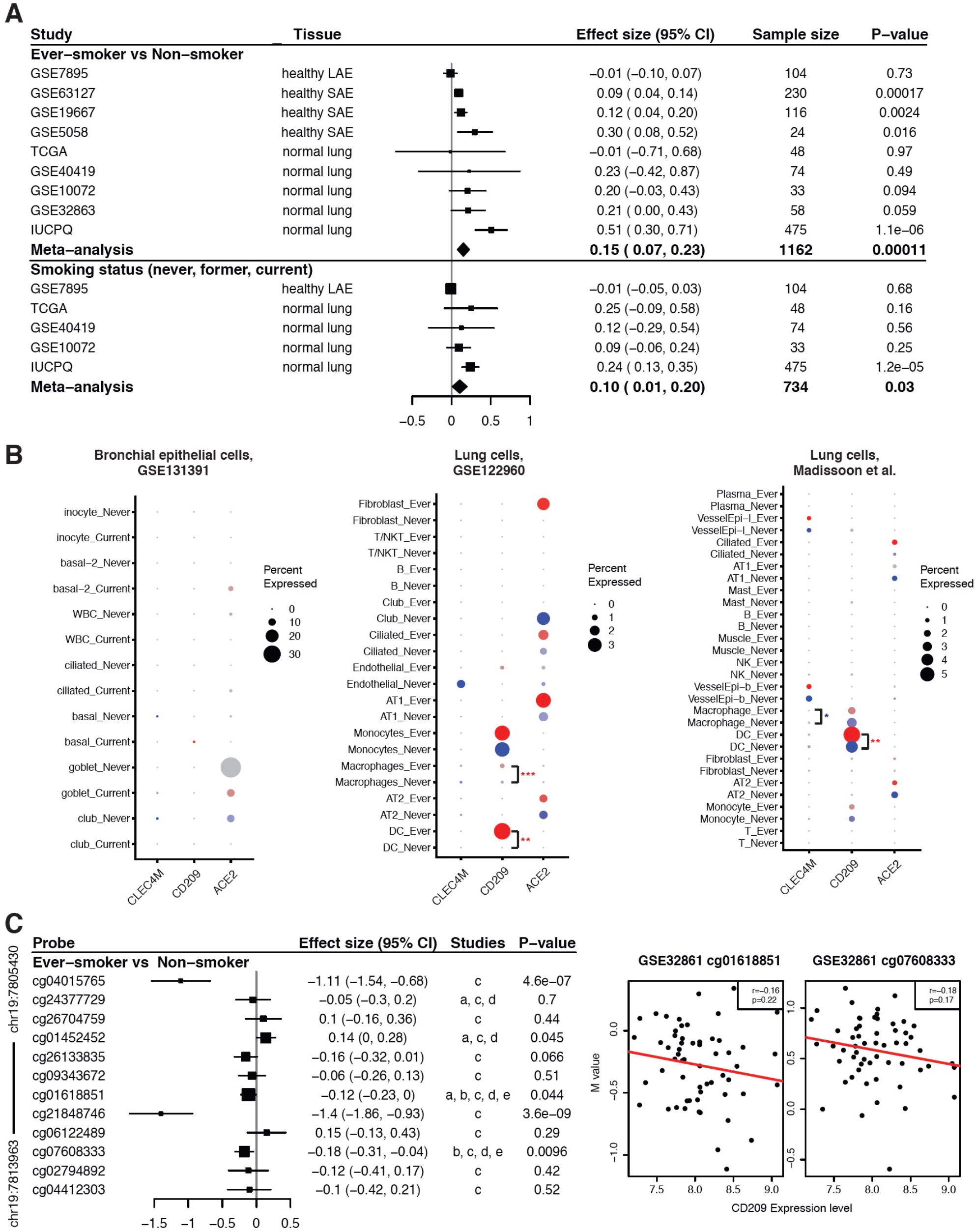
The effect of smoking on pulmonary gene expression of three SARS-CoV-2 receptors. (A) Forest plots for the effects of smoking on *CD209* pulmonary gene expression. The top panel shows a comparison of ever-smokers (including current and former smokers) and nonsmokers, and the bottom panel shows the association of *CD209* gene expression with smoking status (never, former, or current smoker). For each study, pooled effect size, 95% CI and P values were calculated by meta-analysis. The size of the squares is proportional to the weights, which were estimated by the standard “inverse-variance” method for random-effects models in meta-analysis. (B) The expression profiles of *ACE2, CD209* and *CLEC4M* genes in airways of ever-smokers and non-smokers. For each gene, detection rate (showed by dot size, larger dot indicates higher detection rate) and expression level (showed by dot color which is status specific, darker indicates higher expression level) are shown in identified cell types. The goblet cell in never smokers in the GSE122960 dataset was shown in grey because only a limited number of cells was detected. Significance: * P<0.05; ** P<0.01; *** P<0.01. Red * indicates higher expression and blue * indicates lower expression in ever-smokers compared to never-smokers. (C) The association of lung *CD209* methylation with smoking and gene expression. The left panel shows a forest plot for the effects of smoking on *CD209* related methylation probes in normal lung tissues. For each probe, pooled effect size, 95% CI and P values were calculated by meta-analysis from available studies, including a: TCGA, b: GSE32861, c: GSE133062, d: GSE92511 and e: GSE139032. The right panel shows the correlation between *CD209* gene expression levels and measurements of methylation probes (cg01618851, cg07608333) in normal lung tissues from the GSE32861 study.

We next investigated the dysregulation of *ACE2, CD209* and *CLEC4M* in specific types of cells in normal airway samples of never-smokers and ever-smokers. Cell types were identified from scRNA-seq datasets of bronchial epithelium (GSE131391) and lung tissues (GSE122960 and Madissoon) using known markers (Fig. S4, Fig. S5). We found that *ACE2* was mainly expressed in pneumocytes, secretory cells, ciliated cells, never-smokers’ club cells and smokers’ goblet cells (Fig. 2B), which was consistent with the recent study of Ziegler et al.^23^ and our pervious observation^18^. Compared to never-smokers, we found higher detection rate of *ACE2* in ciliated cells in ever-smokers in both lung datasets. The GSE122960 dataset also showed higher expression of *ACE2* in AT1 and fibroblast cells by smoking. Active expression of *CD209* was found in lung DCs, Mø and monocytes (Fig. 2B). Compared to non-smokers, ever-smokers showed increased *CD209* expression in lung DCs (GSE122960 P < 0.01 and Madissoon P < 0.01). Higher CD209 expression was also found in Mø of ever-smokers in the dataset of GSE122960 (P < 0.001). The expression of *CLEC4M* was only notably detected in lung vessel epithelial cells and no significant smoking effect was observed.

Further, we investigated smoking effect on gene methylation of *CD209 and ACE2* by comparing DNA methylation data from smoker and non-smoker lungs in 5 studies (N = 242). Adjusted by age and sex, an overall decrease of methylation was detected across CD209 in smokers, including two significant loci (cg01618851, cg07608333) by meta-analysis and two significant loci (cg04015765, cg21848746) in the GSE133062 study (Fig. 2C). Although not significant with limited sample size, loci cg01618851 and cg07608333 showed trends of negative correlation between *CD209* gene expression and methylation (Fig. 2C). No data was available to investigate the expression-methylation association for loci cg04015765 and cg21848746. In addition, no significant association was observed in *ACE2* (Fig. S6).

### Smoking modulates the function of lung DCs

To verify whether smoking affects the composition of immune cells in lung, we assessed immune cell proportions in normal lung tissues from the above 8 independent bulk transcriptomics studies and evaluated their associations with smoking by meta-analysis. Among 22 immune cell types, we found that resting DCs, CD8^+^ T cells, and activated CD4m T cells were augmented while resting CD4m T cells, activated NK cells, M1 Mø, eosinophils and neutrophils were depleted in lungs of smokers (Fig. 3A left). A similar pattern of associations with smoking history (never, former or current smoker) was observed, with significant positive correlation for CD8 T cells and follicular helper T cells, and negative correlation for CD4 memory resting T cells, activated NK cells, eosinophils and neutrophils (Fig. 3A right). In addition, both resting and activated DCs showed trends of positive association with the smoking history. A trend of increase of DC proportion in smokers was also found in scRNA-seq datasets (Fig. S7).

**Figure 3.**
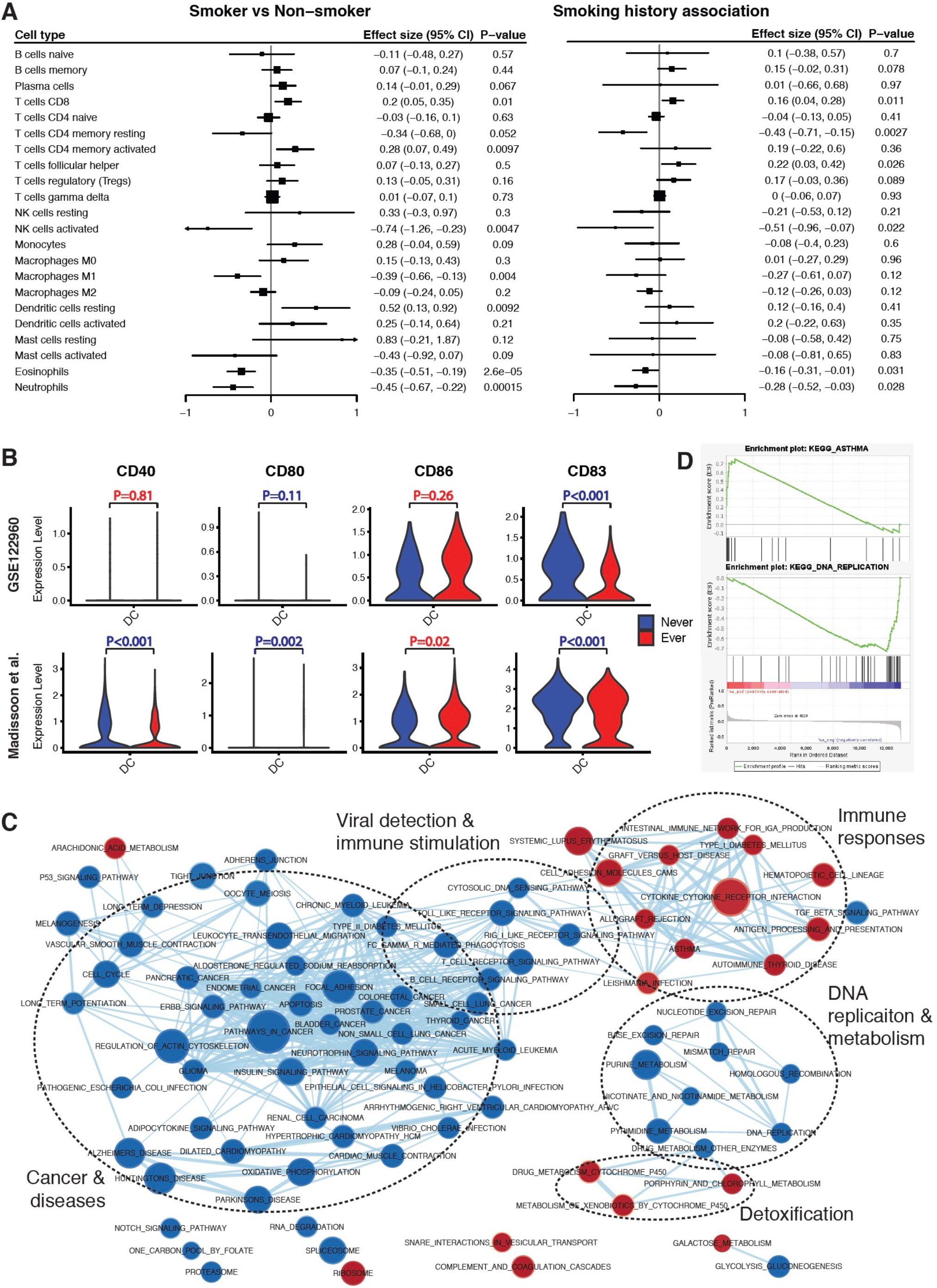
The effect of smoking on lung DCs. (A) Forest plots for the effects of smoking on immune cell composition. For each cell type, the left panel shows a comparison of ever-smoker (including current and former smokers) and nonsmoker groups, and the right panel shows the association of the percentage of the cell in 22 immune cells with smoking status (never, former, or current smoker). For each study, the estimated effect size and 95% confidence intervals (CIs) are plotted. The size of the squares is proportional to the weights, which were estimated by the standard “inverse-variance” method for random-effects models in meta-analysis. (B) Violin plots of the expression of functional markers in DCs from two scRNA-seq datasets. Red P values indicates higher expression and blue P values indicates lower expression in ever-smokers compared to never-smokers. (C) Gene expression networks dysregulated in ever-smoker lungs. The color of a node corresponds to the significance of the gene set (red corresponds to positive effect; blue corresponds to negative effect). Node size corresponds to the number of genes within the gene set. Edge weight corresponds to the number of genes found in both connected gene sets. (D) GSEA enrichment scores of KEGG Asthma and DNA replication pathways. The top portion of the plot shows the running Enrichment Score (ES) for the gene set, the middle portion shows the ranks of the members of the gene set in the ranked list of genes transcriptome wide, and the bottom portion shows the value of the ranking metric in ordered dataset.

Further, we investigated whether smoking affects the maturation of DCs by examining the gene expression of the maturation marker CD83 and the T-cell stimulatory molecules CD86, CD80 and CD40 in the GSE122960 and Madissoon scRNA-seq datasets (Fig. 3B). In DCs of ever-smoker lungs, we found a significant downregulation of *CD83* in both datasets. In the Madissoon dataset, we also found significantly lowered expression of *CD40* and *CD80*. These observations indicate that smoking inhibits both maturation and T cell stimulation ability of DCs, although *CD86* was upregulated in the Madissoon dataset. Pathway analysis also showed that smoking inhibited viral detection and DC maturation including signaling pathways of T cell receptor, B cell receptor, Toll-like receptor (TLR) and RIG-I-like receptor (RLR), as well as pathways involved in Vibrio cholerae and Escherichia coli infections (Fig. 3C). Interestingly, smoker lung DCs also showed upregulated pathways which are involved in immune responses, including antigen processing and presentation, cytokine-cytokine receptor interaction, asthma (Fig. 3D), graft versus host disease and others. In addition, smoking inhibited DNA replication (Fig. 3D), cell cycle, metabolism and DNA repair related pathways, and invoked detoxification processes.

### SARS-CoV-2 modulates the function of DCs and CD209 expression

We next systematically investigated the cell type-specific expression of *CD209* in upper airway, lower airway and peripheral blood by analyzing scRNA-seq datasets from one nasopharynx/pharynx, one BALF and two PBMC (Lee et al., Wilk et al.) studies from COVID-19 patients and healthy controls. Cell types were identified in each dataset (Fig. S8), and the detection rates and average expression of *ACE2, CD209* and *CLEC4M* were compared between groups (Fig. 4A). Compared to healthy controls, patients with mild and severe disease showed higher detection rates of *ACE2* in the nasopharynx/pharynx secretory, ciliated, and pulmonary epithelial cells. No notable expression of *CLEC4M* was observed in any cell type in any of the four datasets.

**Figure 4.**
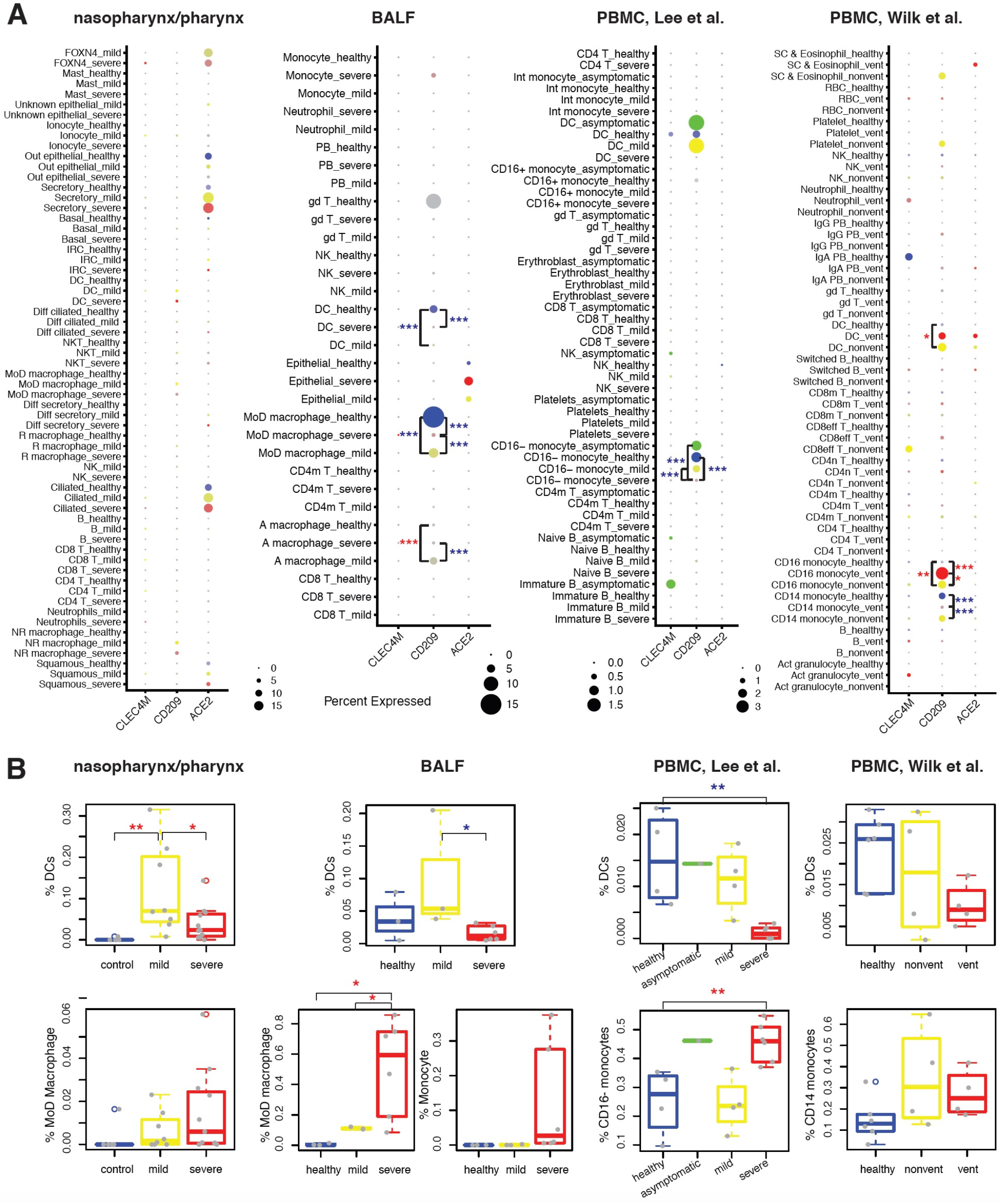
Gene expression differences of three SARS-Cov-2 receptors in upper airway, lung and blood of healthy and COVID-19 cases. (A) Expression profiles of *ACE2, CD209* and *CLEC4M* in cell types identified from four scRNA-seq datasets of nasopharynx/pharynx, BALF and PBMC samples from COVID-19 patients and healthy controls. For each dataset, the detection rate (showed by dot size, larger dot indicates higher detection rate) and expression level (showed by dot color which is status specific, darker indicates higher expression level) of each receptor in identified cell types are shown. The gd T cell in never smokers in the GSE122960 dataset was shown in grey because only a limited number of cells was detected. (B) The proportions of DCs, macrophages or monocytes in WBC in samples from each participant. The differences of expression in DCs, monocytes and monocytes were tested in groups. Significance: * P<0.05; ** P<0.01; *** P<0.01. Red * indicates higher expression and blue * indicates lower expression in severe cases compared to mild cases, in severe cases compared to healthy controls, in mild cases compared to healthy controls, in severe cases compared to asymptomatic cases, in mild cases compared to asymptomatic cases, or in asymptomatic cases compared to healthy controls.

Notable expression of CD209 was found in DCs, airway Mø and blood monocytes (Fig. 4A). Significantly, we found that *CD209* expression in lung monocyte-derived macrophages (MoD Mø) and blood classical (CD16^-^ or CD14^+^) monocytes was severity-dependently decreased in COVID-19, while CD16^+^ monocytes showed a severity-dependent increase of *CD209* expression in the dataset of Wilk et al. In DCs, *CD209* expression was decreased in lungs but increased in blood of COVID-19 cases, as compared to healthy controls. With a small number of detected DCs (N = 12), the Lee PBMC dataset didn’t detect such increase in severe COVID-19 cases. The discrepancy among anatomical sites were also found in the alteration of the DC proportion among white blood cells (WBC) (Fig. 4B). In airway samples, the proportion of DCs was increased in mild cases and decreased in severe cases. A decrease was also found in severe cases in blood, but no notable change was observed in mild cases. Inversely to DCs, the proportions of MoD Mø and monocytes were increased in severe cases in all three tissues. These observations may indicate a site-specific impact of COVID-19 on the differentiation of monocytes to DCs and Mø.

We further identified transcriptome-wide differently expressed genes (DEGs) and signaling pathways in DCs in COVID-19 lung and blood (Fig. 5). Compared to healthy controls, an extensive transcriptomic change was found in lung, with expression of 184 DEGs (93 for mild; 91 for severe, Fig. 5A) changed more than 2-fold, whereas only 15 (8 for mild; 11 for severe, Fig. 5B) were detected in blood. Relatedly, we also found a broad COVID-19 severity-associated activation of viral detection and immune stimulation (cytokine-cytokine receptor interaction, cytosolic DNA sensing, TLR, Nod-like receptor (NLR), chemokine signaling, B cell receptor signaling and Mark signaling) in lung, while only mild immune stimulation (cytokine-cytokine receptor interaction and cytosolic DNA sensing) was found in blood. In both lung and blood, immune response related pathways (graft versus host disease, viral myocarditis, Leishmania infection and others) were activated in mild cases as compared to healthy control but interestingly were inhibited in severe cases as compared to mild cases. Such pattern was also observed on antigen processing and presentation, host metabolism as well as DNA replication in lung but not in blood. Correspondingly, immune initiation related genes (including *CD1A, CD1C, CD1E, CCL17, FCER1A, FCGR2B* and others) and ribosome genes were significantly downregulated in severe COVID-19 cases as compared to healthy controls, whereas inflammatory signals (including interferon genes, *CCL2, CCL4, CCL8* and others) were upregulated (Fig. 5C). Compared to mild cases, chemokines (including *CCL2, CCL4, CCL8*) and MAPK substrates (including *JUN, USF1, FOS*) were upregulated in severe cases, whereas antigen presentation related genes (*HLA-DQA2, C1QC* and *C1QA*) were downregulated. Interestingly in blood, we also found neural disease related pathways were severity-dependently activated and cancer related pathways were inhibited in severe cases (Fig. 5B), which may link to the neurologic and other clinical manifestations of COVID-19.

**Figure 5.**
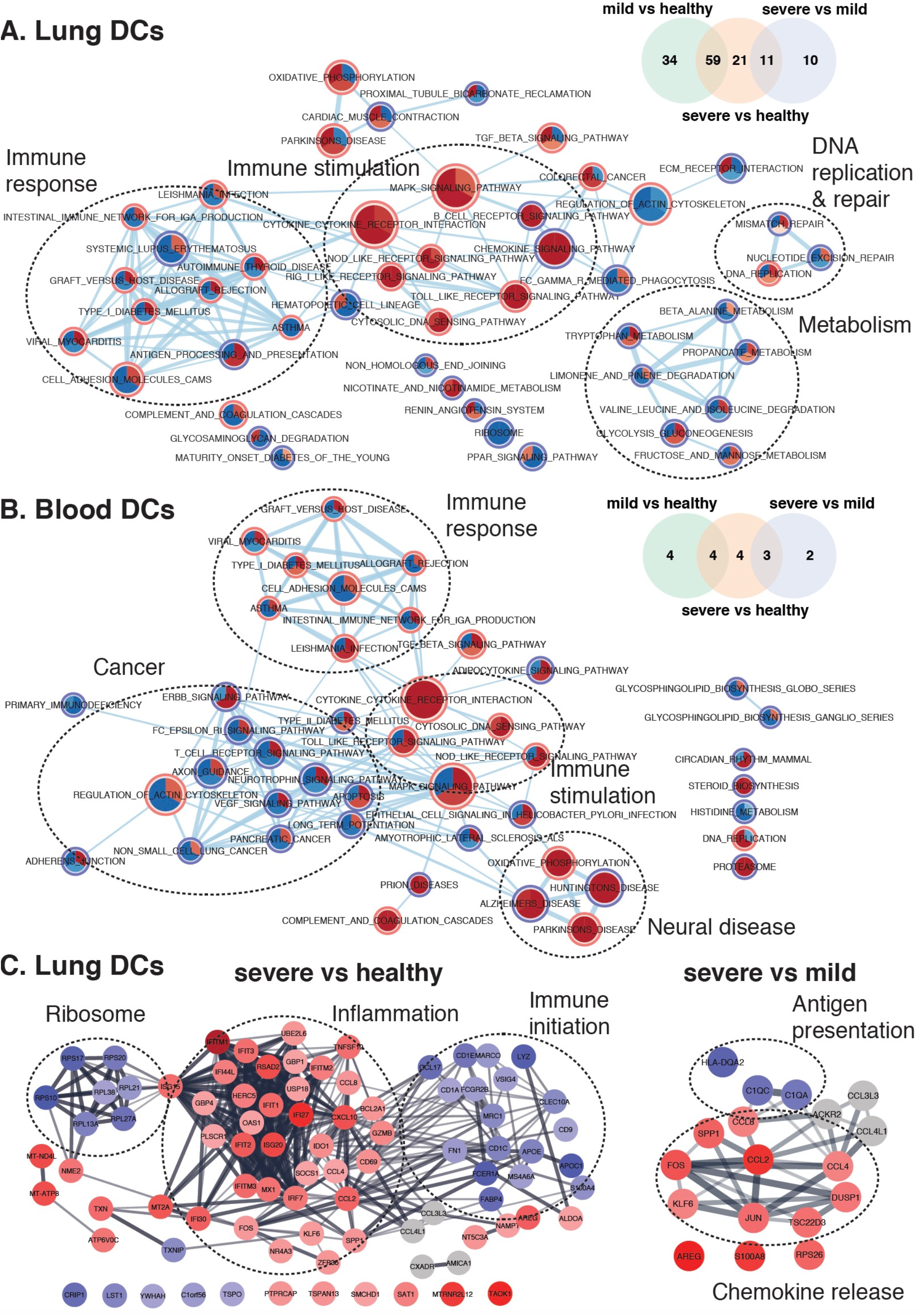
Dysregulated gene expression networks in lung and blood with COVID-19. Each node in network is a pie plot showing three comparisons, including mild vs healthy (top right), severe vs healthy (bottom), and severe vs mild (top left). Node size corresponds to the number of genes in dataset within the gene set. Color intensity inner the node corresponds to the significance of the gene set for this dataset (red is for positive effect; blue is for negative effect). Orange outer circles show the overlapping gene sets found in (A) lung DCs and (B) blood DCs, while blue outer circles show the uniquely detected gene sets. Edge weight corresponds to the number of genes found in both connected gene sets. Venn diagrams show the distribution of significant altered genes more than 2 folds detected from three comparisons in each dataset. (C) shows STRING networks of changed genes in lung DCs in severe COVID-19 cases compared to healthy (left) and mild cases (right). The intensity of filling color corresponds to the log_2_FC of the gene (red is for positive effect; blue is for negative effect).

The expression of the maturation marker (*CD83*), T-cell stimulation molecules (*CD40, CD80, CD86*) and antigen presentation molecules (*HLA-DQA2* and *FCER1A*) showed the dysregulation of DC function in COVID-19 (Fig. 6A). In lung, *CD40, CD80, CD83* and *HLA-DQA2* were upregulated while *CD86 and FCER1A* were decreased in mild COVID-19 as compared to healthy controls. Similar patterns were observed on *CD40, CD86* and *HLA-DQA2* in both blood datasets while inconsistent results of *CD83* and *FCER1A* were observed in Lee and Wilk PBMC datasets. Compare to mild cases, severe COVID-19 showed upregulated *CD40* as well as downregulated *CD83, HLA-DQA2* and *FCER1A* in all three tissues. Interestingly, asymptomatic cases showed similar expression of those markers with healthy controls in blood.

**Fig 6.**
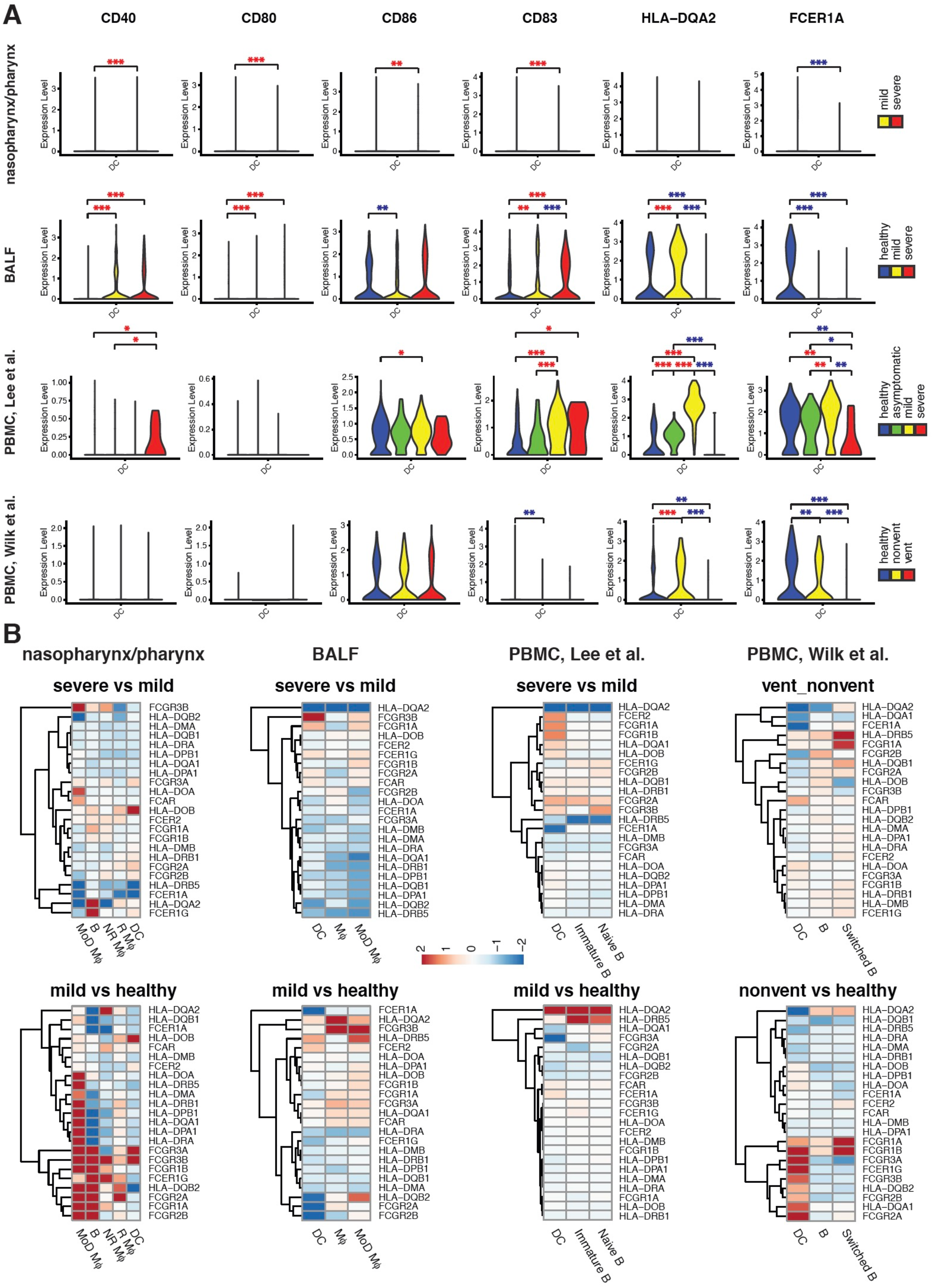
The expression profiles of functional markers in DCs at three sites. (A) The expression disparity of markers of maturation (CD83), T-cell stimulation molecules (CD40, CD80, CD86) and antigen presentation molecules (HLA-DQA2 and FCER1A) in DCs of COVID-19 cases and healthy controls. 1 upper airway (nasopharynx/pharynx), 1 lung (BALF) and 2 blood (PBMC) scRNA-seq datasets were analyzed. Significance: * P<0.05; ** P<0.01; *** P<0.01. Red * indicates higher expression and blue * indicates lower expression in severe cases compared to mild cases, in severe cases compared to healthy controls, in mild cases compared to healthy controls, in severe cases compared to asymptomatic cases, in mild cases compared to asymptomatic cases, or in asymptomatic cases compared to healthy controls. Despite interest, we did not compare COVID-19 nasopharynx/pharynx samples to healthy controls due to the limited detection of DCs (N = 1). (B) The dysregulation of gene expression of MHCII and FcRs in APCs at 3 sites with COVID-19. For each gene, the alteration of gene expression in APCs of severe COVID-19 cases compared to mild cases, and that in mild COVID-19 cases compared to healthy controls are visualized in heatmaps. Red indicates upregulation and blue indicates downregulation. The scale corresponds to log_2_FC.

In addition, we investigated the expression of MHC class II molecules (MHCII) and Fc receptors (FcRs) and found that they were broadly activated in APCs in lungs of mild COVID-19 patients, with a MHCII subset (*HLA-DRA, HLA-DMB, HLA-DRB1, HLA-DPB1, HLA-DQB1, HLA-DMA*) were downregulated (Fig. 6B). Compared to mild cases, severe cases showed decreased MHCII expression in airway APCs but not significantly in blood. The result was in agreement with above pathway analysis that antigen processing and presentation were activated in mild cases but inhibited specifically in lungs of severe cases. Significantly in lung and blood APCs, HLA-DQA2 expression were increased in mild cases as compared to healthy controls but downregulated in severe cases as compared to mild cases. Interestingly, compared to healthy controls, the global change of MHCII and FcRs gene expression in blood were associated with COVID-19 severity, with upregulation in asymptomatic cases, no significant change in mild cases and downregulation in severe cases (Fig. S9).

Together, these observations indicate the ability of DCs for T-cell stimulation is induced but the maturation of DCs is inhibited in both lung and blood of severe COVID-19, and specifically the antigen presentation ability of APCs is reduced in lung, which is the primary site of infection.

### Conventional DC subsets were depleted and new immature DC subsets were accumulated in severe COVID-19

We further investigated the variation of DC subsets in nasopharynx/pharynx, BALF and PBMC samples from COVID-19 patients and healthy controls. Based on known markers (Fig. S10), four DC subsets were identified in the nasopharynx/pharynx samples, including type I and II conventional dendritic cells (cDC1 and cDC2), Mø-like DC which expressed Mø markers S100A8 and S100A9, Plasmacytoid dendritic cell (pDC) and a new subset of CD22^+^ITM2C^+^ DC; seven subsets were identified in the BALF samples, including cDC1, cDC2, mature cDC2, Mø-like DC, pDC and new subsets CD22^+^ DC and ANXA1^+^ DC; four subsets were identified in the PBMC samples from the Lee dataset, including cDC1, cDC2, mature cDC2 and Mø-like DC; four subsets were identified in the PBMC samples from the Wilk dataset, including cDC1, cDC2, mature cDC2 and Mø-like DC (Fig. 7A). The integration of all datasets found that these subsets were conservative among tissues (Fig. S11). CD22^+^ITM2C^+^ DC in nasopharynx/pharynx samples were clustered together with CD22^+^ DC in BALF samples, which were characterized by the activated immune response (*q* = 1.17 × 10^−5^) and interferon gamma response (*q* = 1.17 × 10^−5^).

**Fig 7.**
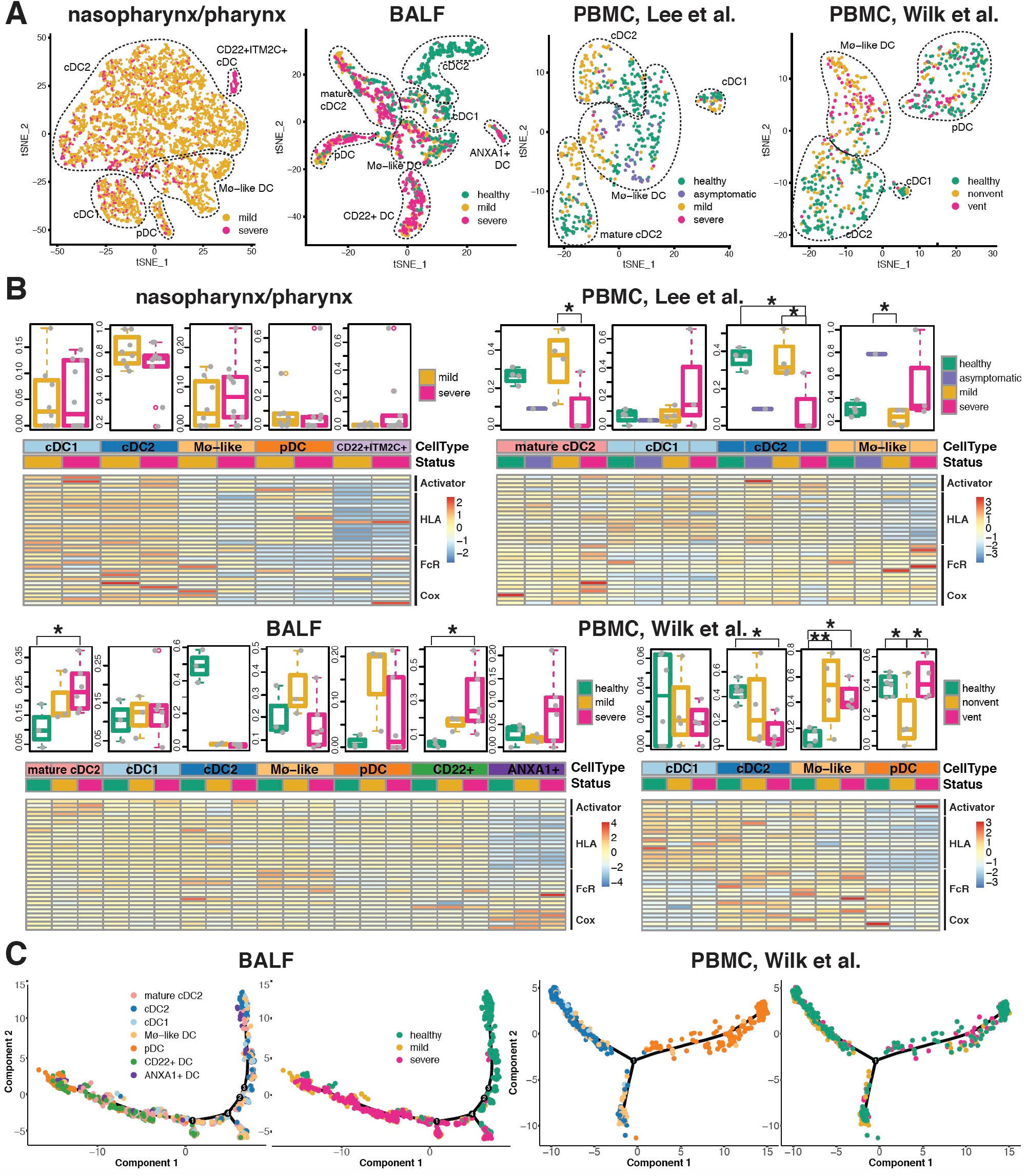
The variation of DC subsets in COVID-19 samples and healthy controls at three sites. (A) DC subsets identified in single-cell transcriptome profiles from COVID-19 mild and severe cases and healthy controls. 1 upper airway (nasopharynx/pharynx), 1 lung (BALF) and 2 blood (PBMC) scRNA-seq datasets were analyzed. (B) The proportions of DC subsets in samples from each participant was shown in the top panel for each dataset. The differences of proportion of each DC subset were tested in groups. Significance: * P<0.05; ** P<0.01. The bottom panel shows gene expression profile of the T-cell stimulation activators (CD83, CD40, CD80, CD86), MHCII and FcRs in subsets in samples with different disease status. Red indicates upregulation and blue indicates downregulation. The scale corresponds to the log_2_ (normalized scRNA-seq read count) which was centered and scaled by row. (C) Single-cell trajectory of DCs inferred from scRNA-seq data from BALF and PBMC samples. For each dataset, the right shows ordered cells from COVID-19 mild and severe cases, and healthy controls. The left shows ordered cells labeled with identified cell types.

Among DCs, the proportion of cDC2 was severity-dependently decreased in both lung and blood of COVID-19 (Fig. 7B). Inversely, the mature cDC2 was increasingly correlated with disease severity in lung BALF samples. Such increase was also found in PBMC with mild COVID-19 but not in severe cases which might be due to its limited cell detection of DC cells (N=12). pDC showed a notable augmentation in BALF and depletion in PBMC in mild cases but not in severe cases, which may reflect the different migration activeness of pDC from blood to lung in mild and severe COVID-19. Interestingly, all tissues in severe cases showed augmentation of immature DC subsets with decreased mature markers *CD83*, T-cell stimulation activators (*CD40, CD80, CD86*) and HLA genes, including CD22^+^ITM2C^+^ DC and Mø-like DC in nasopharynx/pharynx samples, ANXA1^+^ DC and pDC in BALF samples, Mø-like DC and pDC in PBMC samples. Indeed, in nasopharynx/pharynx samples, the proportion of CD22^+^ITM2C^+^ DC was significantly positively correlated with that of M0 Mø (*P* = 8.6 × 10^−5^), resting CD4 memory T cells (*P* = 0.028) and resting mast cells (*P* = 0.024) (Fig. S12). In addition, we found the BALF specific ANXA1^+^ DC subset showed high expression of COX related genes *PTGS1, PTGS2, HPGDS, ALOX5* and *LTC4S*, which may be responsible for the DC immunity suppression^24^, together with *ANXA1*^*25*^.

We observed a clear shift from conventional DCs to pDC on single cell trajectory in COVID-19 BALF and PBMC, in response to viral infection (Fig. 7C). However, the shift was trapped in the path in severe cases, producing immature subsets. The blockade was also observed in nasopharynx/pharynx with severe COVID-19 (Fig. S13). Further we found that both bursting frequency and size of MCHII genes were inhibited in the lung specific immature subset ANXA1^+^ DC as compared to cDC2 and pDC (Fig. S14). Increased frequency and size of Cox genes were also observed. This indicate the SARS-CoV-2 infection may inhibit the DC maturation by regulating the expression kinetics parameters (bursting frequency and bursting size) of related genes.

## Discussion

ACE2, DC-SIGN and L-SIGN are three receptors of both SARS-CoV and SARS-CoV-2. No data is available to indicate they are dependent viral entry portals and we did not find significant correlations of the gene expression between ACE2 and that of DC-SIGN and L-SIGN (Fig. S15). Given that DC-SIGN is mainly expressed on the surface of DCs and Mø, the viral infection to immune cells may directly link to the immunopathogenesis and disease severity of COVD-19.

Indeed, we found higher gene expression of *ACE2* and *CD209* in upper airway cells of COVID-19 ARIs compared to non-viral ARIs, and *CD209* plasma protein expression was correlated COVID-19 risk and severity. We also observed their association with the variation of immune cell activation in upper airway cells of COVID-19 ARIs that high expression was only found in the T/Mø activated cells. This might be a result from the immune stimulation by intensive viral attack to ACE2 and DC-SIGN expressing cells. This may also reflect the negative correlation of *CD209* expression with neutrophil percentage and the positive correlation with T cell percentage, which were detected by our large-scale population genetic association study. Indeed, the activated neutrophils produce TNF-alpha and induce DC maturation which lower the *CD209* expression^26^; *CD209* mediates the activation of T cells^27^. The neutrophil-to-lymphocyte ratio has been studied as a severity marker in COVID-19^28^ and as a valid prognostic factor in various solid tumors and other chronic diseases^29^. Therefore, the interaction of *CD209* and DCs with neutrophils and T cells may play an important role in mediating cytokine storm coupled with limited anti-viral capability, which are keys in COVID-19 pathogenesis^30^. Further investigation is required to elucidate this interaction and the causal effect of *ACE2* or *CD209* to the COVID-19 risk.

For the first time, we performed a systematic investigation of the expression of SARS-CoV-2 receptors in single cells of upper airway, lower airway and blood of COVID-19 patients. We found *CD209* expression and the percentage of DCs and MoD Mø or monocytes were associated with COVID-19 severity. Interestingly, the associations were site specific. In the lung and upper airway with mild infection, DCs were induced for an active immune response and an cell augmentation. In the cases of severe infection, the decrease of DCs proportion and maturation could be a sign exhaustion of DCs. Also, the decreased expression of *CD209* in DCs of COVID-19 lungs may be a result of cell death caused by direct viral infection. Differently, blood showed induced DC activation and increased *CD209* expression, which may indicate that DCs are stimulated in blood but not attacked by virus. Indeed, no SARS-CoV-2 sequencing reads were detected from COVID-19 PMBC^16^. Interestingly, DCs in lungs of severe COVID-19 showed upregulation of T cell stimulation but downregulation of antigen presentation and maturation, indicating that DCs may stimulate innate immune system but fail to recognize SARS-CoV-2. This may lead to the negatively correlated cytotoxic follicular helper response with antibody levels to SARS-CoV-2 spike protein in hospitalized COVID-19 patients^31^. SARS-CoV-2 might use an escape mechanism similar to HIV-1 which hides in DCs and uses DC-SIGN as a trans-receptor for efficient transmission to T cells, and also similar to *Mycobacteria tuberculosis (M. tb)* which subverts DCs by targeting DC-SIGN and inhibiting maturation^32^. The stimulated and “blinded” immune systems could be responsible for the cytokine storm as well as severe ARDS suffering COVID-19 critical cases. MHCII genes especially HLA-DQA2 might be involved in this pathogenesis, which were found to be decreased in lung of severe cases compared to mild cases. This may also link to the olfactory impairment which is often seen in patients with COVID-19, given that MHC are essential in individual olfactory perception^33^.

The observed broad downregulation of MHCII genes in APCs was aligned with recent findings that dysfunctional monocytes^34,35^ and B cells^36^ in severe COVID-19. Here, we report a shift from cDCs to pDCs in COVID-19 cases, which leads to the marked disease severity-dependent depletion of cDC2 in all studied tissues. Strikingly, the shift was blocked in immature subsets including CD22^+^ DC and ANXA1^+^ DC with suppressed activation markers. This immunosuppression of DCs may contribute to the variation of disease severity through the interactions between DCs and T cells and other immune cells. Indeed, we found the immature subtype CD22^+^ITM2C^+^ DC was significantly positively correlated with the resting subsets of Mø, CD4mT and mast cells in upper airway.

In this study, we also investigated the smoking effect on respiratory gene expression of these three receptors of both SARS-CoV and SARS-CoV-2. We found ever-smokers had significantly higher *ACE2* and *CD209* gene expression than non-smokers, respectively in lung AT1 cells and monocytes/DCs. Our results indicate that this smoking effect might be mediated by methylation regulation. Indeed, methylation switch *CD209* genes during the differentiation of human Monocytes to dendritic cells^37^. Also, DCs maturation and signaling pathways such as TLR signaling for viral detection and innate immune initiation were inhibited by smoking. Thus, smoking may accomplice the escape of SARS-CoV-2 from immune surveillance by targeting DCs, as shown in this study that antigen presentation and maturation are inhibited in DCs in severe COVID-19 lungs. Indeed, cigarette smoking has been confirmed to increase the susceptibility to various respiratory diseases such as pneumococcal disease and pneumonia^38,39^. Severance et al. also identified smoking status as a risk factor for the exposure of four non-SARS coronavirus strains among US population^40^. Interestingly, we also found smoking induces the DC proportion and enhances immune responses related pathways in DCs. These results indicate that DCs from smoking individuals have lower ability to stimulate T cells to attack a pathogen but actively induce immune responses and release cytokines, which may partially contribute to the cytokine storm in severe COVID-19 patients. Together with the upregulation of *ACE2* and *CD209* expression by smoking, this may explain the clinical presentation of 1,099 cases of COVID-19 that the smokers had a higher risk of severe disease^41^.

Collectively, this study provides important pieces of puzzle of COVID-19 about dysfunctional DC cells from the expression of receptors of SARS coronavirus, the effect and potential risk of smoking, and remodeling of DC activation and redistribution of DC subsets. Although further studies with a larger sample size of single cell or single cell type studies will be needed to properly assess this matter, this study provide important hints for vaccine development, as well as the variation of COVID-19 severity and pathogenesis.

## STAR★Methods

### Key Resources Table

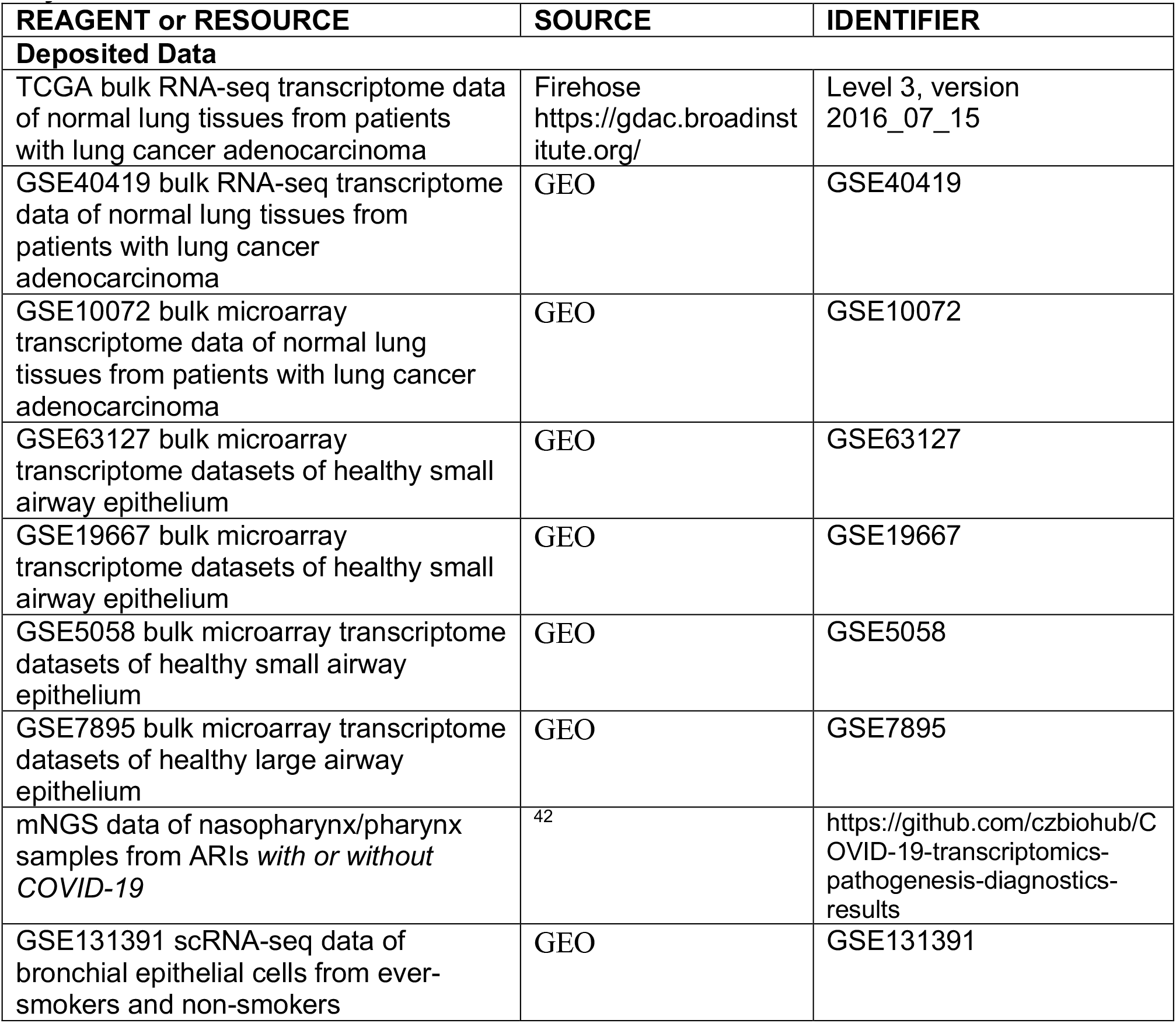
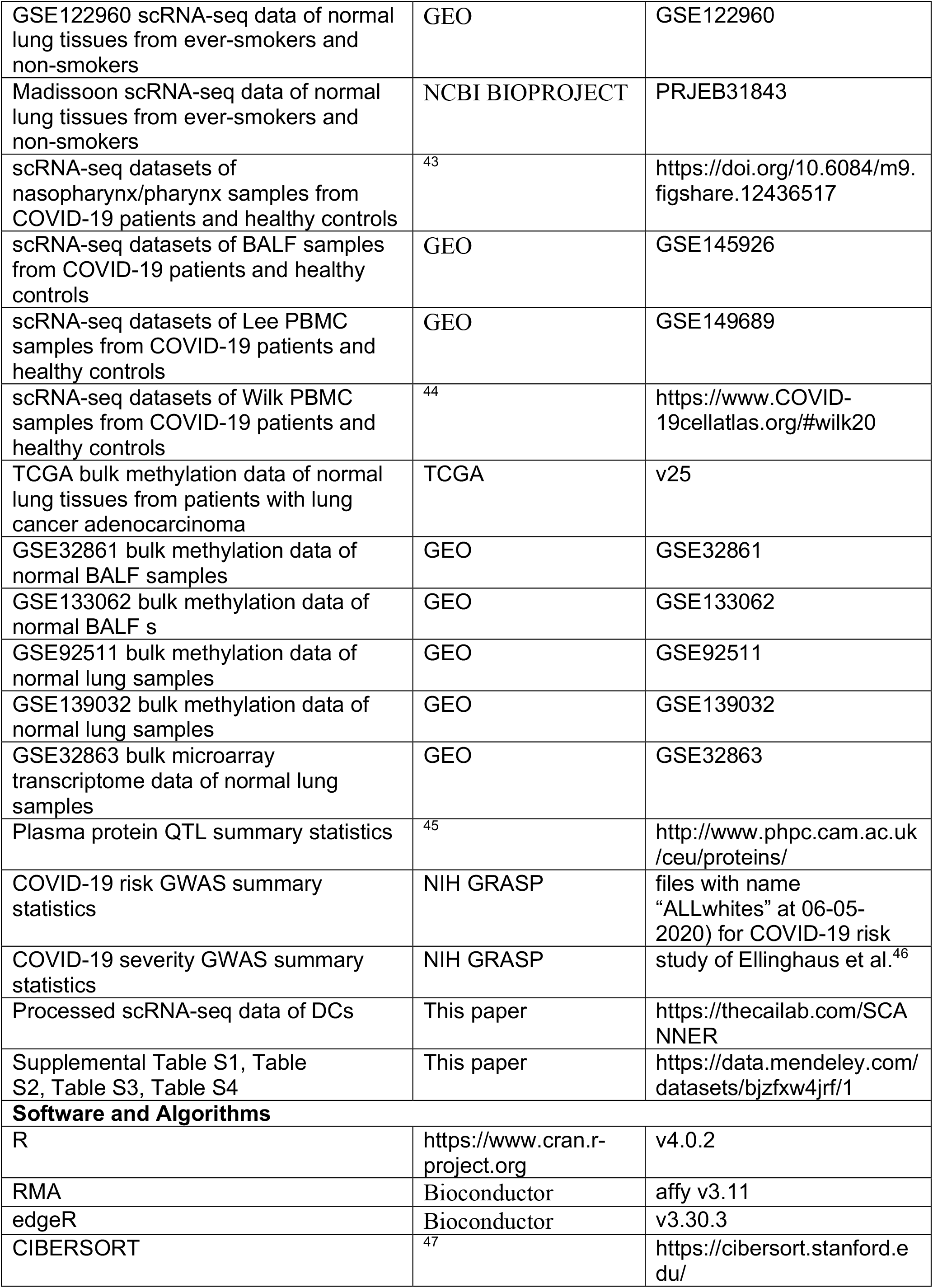
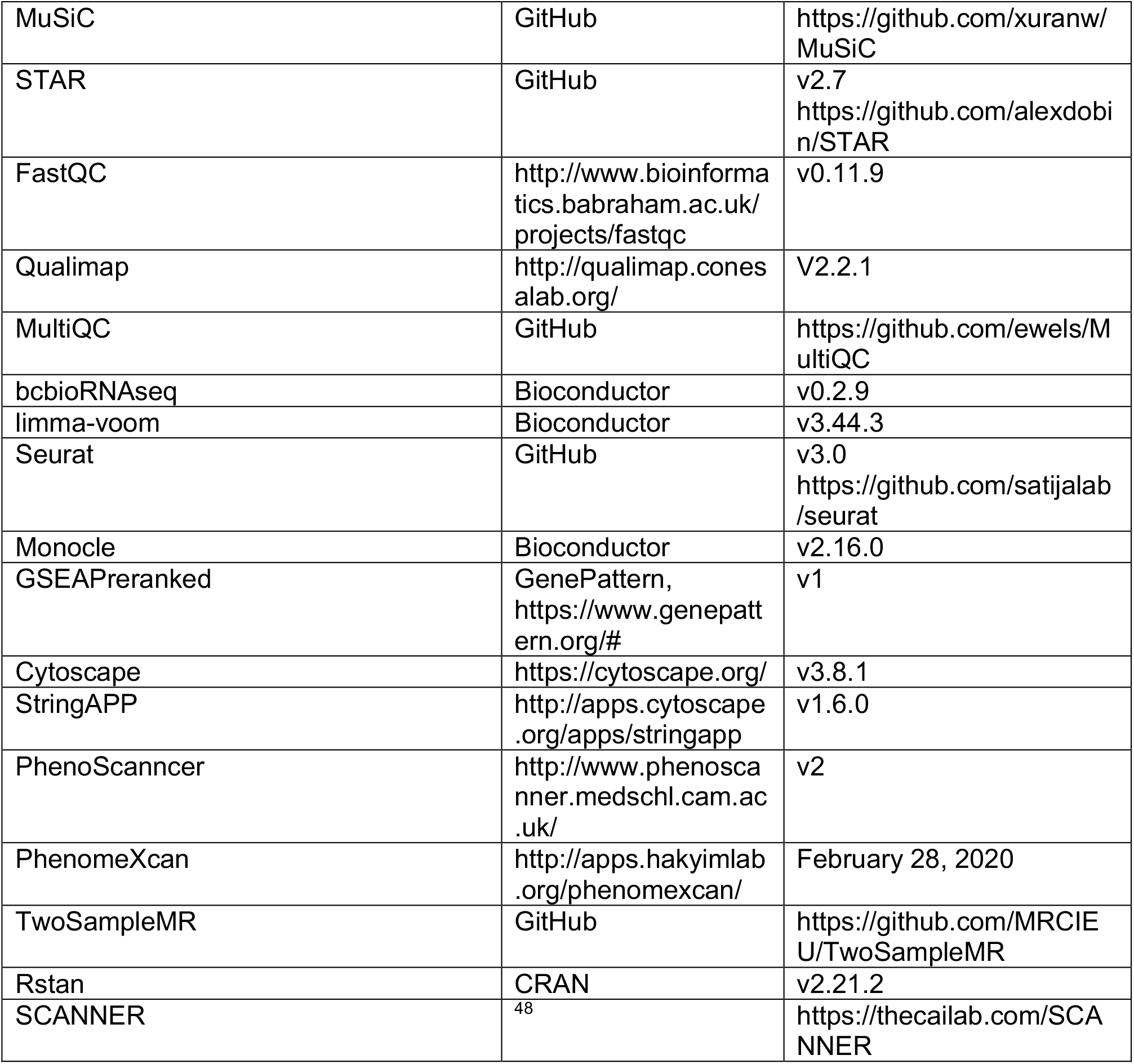

### Bulk transcriptomics

#### Normal lung and airway samples from ever-smokers and non-smokers

We used eight datasets (N = 1162) to investigate the gene expression of three SARS coronavirus receptors (ACE2, DC-SIGN and L-SIGN, with gene symbols *ACE2, CD209* and *CLEC4M*) in bulk tissues, including four datasets of normal lung tissues from patients with lung cancer adenocarcinoma (TCGA RNA-seq dataset (N = 48)^49^, Institut universitaire de cardiologie et de pneumologie de Québec (IUCPQ) microarray dataset^50^ (N = 475), Gene Expression0020Omnibus (GEO) GSE40419 RNA-seq dataset^51^ (N = 74) and GSE10072 microarray dataset^52^ (N = 33)), three microarray datasets of healthy small airway epithelium (SAE) samples (GSE63127^53^ (N = 230), GSE19667^54^ (N = 116) and GSE5058^55^ (N = 24)) and a microarray dataset of healthy large airway epithelium (LAE) samples from GSE7895^56^ (N = 104). We studied the Reads Per Kilobase per Million mapped reads (RPKM) values for RNA-seq data and Robust Multi-array Average (RMA)^57^ values for microarray data. All data were log2 transformed to improve normality. Never-smokers and ever-smokers (including current and former smokers) were identified in each original study based on self-reported smoking history. Multivariate linear regressions were used to test the association of *ACE2, CD209* and *CLEC4M* gene expression with smoking status adjusted for age and sex. Meta-analysis was performed by pooling effect size and standard error estimated from each study using a random effects model with Hunter-Schmidt estimator.

#### Upper airway samples from patients with or without COVID-19

We studied the metagenomic next generation sequencing (mNGS) data of nasopharynx/pharynx samples from 197 patients with acute respiratory illnesses, including 94 patients confirmed with SARS-CoV-2 infection by clinical PCR and 103 with no virus detected^42^. The host gene expression read counts and the CPM (read counts for million) of SARS-CoV-2 virus were analyzed. We normalized the read counts of *ACE2* and *CD209* using the TMM method and calculated CPM using edgeR^58^. The data for *CLEC4M* was not available.

We deconvoluted the immune cell composition from these bulk RNA-seq data using CIBERSORT^47^ with its signature matrix, LM22. In each sample, the proportions of 22 human hematopoietic cell types were inferred, including naïve and memory B cells, plasma cells, 7 T cell types (CD8, CD4 native, memory resting CD4, memory activated CD4, follicular helper, regulatory (Tregs) and gamma delta), NK cells (resting and activated), monocytes, macrophages (M0, M1 and M2), dendritic cells (resting and activated), mast cells (resting and activated), eosinophils and neutrophils. Using transcriptome profiles of DC subsets identified in below scRNA-seq datasets of nasopharynx/pharynx samples, we also estimated the composition of DC subsets in these bulk RNA-seq data using MuSiC^59^.

Linear regression was used to test the difference of gene expression or immune cell proportions between sample groups as well as their association with SARS-CoV-2 viral load in samples.

#### Blood samples from ARDS cases and controls

From the Molecular Epidemiology of ARDS (MEARDS) prospective cohort study^60^,160 Acute Respiratory Distress Syndrome (ARDS) cases and 142 controls were recruited for RNA-Seq analysis of blood samples. RNA was extracted by PAXgene Blood RNA Kit (QIAGEN) and selected by using oligo(dT) beads. Sequencing libraries were built using MGIEasy RNA Library Prep Kit and subsequently sequenced on the MGISEQ-2000 platform. The 100 bp pair-end sequencing reads were filtered to remove low-quality, adaptor-polluted and high content of unknown base (N) reads. Further, sequencing reads were mapped to reference genome GRCh38 using STAR^61^ and counts of reads aligning to known genes were generated by featureCounts^62^. Quality controls were performed for evenness of coverage, rRNA content, genomic context of alignments using FastQC (http://www.bioinformatics.babraham.ac.uk/projects/fastqc), Qualimap^63^, MultiQC (https://github.com/ewels/MultiQC), and bcbioRNAseq (version 0.2.9)^64^. Then, limma-voom^65^ was used to perform whole-transcriptome differential expression analysis from raw counts.

### Single-cell transcriptomics

#### Normal airway samples from ever-smokers and non-smokers

We analyzed three single-cell RNA sequencing datasets including GSE131391^66^, GSE122960^67^ and a dataset from Madissoon *et al*.^68^. The GSE131391 dataset focused on bronchial epithelial cells, ALCAM+ epithelial cells and CD45+ white blood cells from 6 never- and 6 current-smokers. The GSE122960 dataset was from lung tissues of 8 lung transplant donors, including 5 never- and 3 ever-smokers. The Madissoon dataset was from lung tissues of 4 never- and 3 ever-smokers. Sequencing read counts in single cells were downloaded, and subsequent data analyses were performed using the Seurat 3.0 package^69^, including data normalization, high variable feature selection, data scaling, dimension reduction and cluster identification. The Louvain method was used on the top 20 principle components to identify clusters, which were visualized in reduced dimensions of Uniform Manifold Approximation and Projection (UMAP). Cell types were assigned to clusters based on markers used in each original study. In GSE131391 samples, we identified club cells, goblet cells, ciliated cells, CD45+ white blood cells (WBC), ionocytes, basal cells and a basal cell sub-population (basal-2) which was shown in smokers. In GSE122960 samples, we identified alveolar type I cells (AT1), alveolar type II cells (AT2), macrophages, monocytes, endothelial cells, ciliated cells, club cells, B cells, T/NKT cells, DCs and fibroblast cells. In samples from the study of Madissoon et al., we identified AT1, AT2, macrophages, monocytes, ciliated cells, B cells, T cells, NKT cells, DCs, mast cells, fibroblast cells, muscle cells, plasma cells, blood vessel endothelial cells (VesselEpi-b) and lymph vessel endothelial cells (VesselEpi-l).

#### Upper airway, lower airway and peripheral blood samples from COVID-19 patients and healthy controls

We investigated the gene expression of *ACE2, CD209* and *CLEC4M* in scRNA-seq datasets of nasopharynx/pharynx, BALF and PBMC samples from COVID-19 patients and healthy controls. The nasopharynx/pharynx data were available in the study of Chua et al.^43^ with the count matrix and cell type identification derived from 11 moderate and 8 critical COVID-19 patients and 5 healthy controls. We also downloaded the count data of BALF samples from 6 severe and 3 mild COVID-19 patients as well as 3 healthy controls from GEO GSE145926^15^. Also, the count data from PBMC samples of 6 severe, 4 mild, and 1 asymptomatic COVID-19 patients and 4 healthy controls were downloaded from GEO GSE149689 of Lee et al.^70^. In addition, we analyzed another PBMC scRNA-seq dataset from the study of Wilk et al.^44^, which was from 6 healthy controls and 7 COVID-19 patients. 4 of 8 COVID-19 samples were collected from patients who were diagnosed with ARDS and were ventilated. Two samples were collected from one patient in the non-severe condition without ventilation and in the severe condition with ventilation.

scRNA-seq data processing was performed using Seurat with steps same as above. 110145 cells including 9326 DCs were analyzed in the nasopharynx/pharynx dataset. 71415 cells including 1484 DCs were analyzed in the BALF dataset. 58187 cells including 526 DCs were analyzed in the Lee et al. PBMC dataset. 36925 cells including 514 DCs were analyzed in the Wilk PBMC dataset. Cell types in the nasopharynx/pharynx dataset and the Wilk et al. PBMC dataset were identified by their original studies. Following the approaches used in the study of Wilk et al.^44^, we used SingleR^71^ and markers used in each original study, we identified cell types for cell clusters detected in the BALF and Lee et al. PBMC datasets. In the nasopharynx/pharynx samples, we identified squamous cells, non-resident macrophages (NR macrophages), monocyte-derived macrophages (MoD macrophages), resident macrophages (R macrophages), neutrophils, CD4^+^ T cells, CD8^+^ T cells, B cells, ciliated cells, NK cells, NKT cells, Diff ciliated, DCs, IFNG-responsive cells (IRC), basal, secretory, differentiating secretory (Diff secretory), outliers epithelial cells (Out epithelial), ionocytes, Unknown epithelial cells, mast cells and FOXN4^+^ cells. In the BALF samples, we identified CD8^+^ T cells, CD4^+^ memory T cells (CD4m T), gamma delta T cells (gd T), alveolar macrophages (A macrophages), MoD macrophages, epithelial cells, DCs, NK cells, plasma B cells (PB), neutrophils and monocytes. In the Lee PBMC dataset, we identified immature B cells, naive B cells, CD4m T cells, CD8^+^ T cells, CD4^+^ T cells, erythroblasts, gd T cells, CD16^+^ monocytes, CD16^-^ monocytes, platelets, NK cells, DCs and intermediate monocytes (Int monocytes). In the Wilk PBMC dataset, we identified activated granulocytes (Act granulocytes), B cells, CD14^+^ monocytes, CD16^+^ monocytes, CD4^+^ T cells, CD4m T cells, naive CD4^+^ T cells (CD4n T), effector CD8^+^ T cells (CD8eff T), CD8m T cells, class-switched B cells (Switched B), DCs, gd T cells, IgA-producing PB cells (IgA PB), IgG-producing PB cells (IgG PB), neutrophils, NK cells, DCs, platelets, red blood cells (RBC), as well as stem cells and eosinophils (SC & Eosinophil). To compare gene expression in groups, Wilcoxon rank sum test was used.

To identify DC subsets, we used well evidenced markers for cDC1 (*CLEC9A, C1orf54, HLA-DPA1, CADM1, CAMK2D, XCR1*), cDC2 (*CD1C, FCER1A, CLEC10A*), pDC (*DAB2, GZMB, JCHAIN, SERPINF1, ITM2C, CLEC4C, LILRA4*), mature DC (*LAMP3, CD86, CD83, CD40, ICAM1, ITGA4, CCL8, CCL22, CCL17, CXCL8, CXCL9, CXCL10, CXCL11, CCR7*), macrophage-like DC (*S100A9, S100A9, S100A8, VCAN, LYZ, CD14*), AXL^+^ DC (*ANXA1, AXL, PPP1R14A, SIGLEC6, CD22*) and non-classical DC (*FCGR3A, CX3CR1*). To evaluate the similarity of DC subtypes identified in above scRNA-seq datasets, we integrated these datasets using Seurat and visualized the integrated matrix with UMAP. Monocle^72^ was used to infer the single cell trajectory.

### Methylation analysis of *CD209* and *ACE2*

In this study, five datasets (N = 237) of normal lung tissues were used to investigate the association of smoking and DNA methylation of CD209, including two datasets of normal lung tissues from patients of lung adenocarcinoma from TCGA^49^ (Illumina 450K BeadChip, N = 55) and GSE32861^73^ (Illumina 27K BeadChip, N = 59), one dataset of BALF cells from GSE133062^74^ (Illumina MethylationEPIC BeadChip, N = 35), and two datasets of normal non-malignant lung samples from GSE92511^75^ (Illumina 450K BeadChip, N = 16) and GSE139032^76^ (Illumina 27K BeadChip, N = 77). Smoking history was determined by self-reported history. Multivariate linear regressions were used to test the association of the M-values of *CD209* methylation probes with smoking status adjusted for sex and age. Using the effect size and standard error estimates from each study, a random-effects meta-analysis was performed as described before. With paired data from the GSE32863^73^ mRNA expression and the GSE32861 methylation measurements, we also evaluated the correlation of CD209 expression abundance and methylation level in 58 paired samples.

### Group comparison and association testing

Linear regression was used to evaluate the association between variables and perform two-group comparison. To evaluate the association of *CD209* expression with tumor stage and survival of lung adenocarcinoma tumors, ordinal regression and Cox proportional hazard model were used respectively.

### Pathway and network analyses

Pathway and network analyses were performed to systematically investigate biological processes involved in smoking or COVID-19. We calculated transcriptome-wide fold changes (FC) of expression and ranked the differential expression according to log_2_FC. For Based on these ranks, preranked Gene Set Enrichment Analysis (GSEA)^77^ was applied to evaluate the enrichment of Kyoto Encyclopedia of Genes and Genomes (KEGG)^78^ pathways. Further, the summary statistics of GSEA analysis were used to build connection networks of detected pathways by the Enrichment Map implemented in Cytoscape^79^. Pathways with Q-value<0.2 were included into networks and the connectivity strength was determined by an edge cutoff as 0.25. In addition, networks of genes with absolute value of the average of log_2_FC >1 were constructed using the Cytoscape StringAPP^80^.

### Inference and comparison of transcriptional bursting kinetics between DC subsets

To infer the bursting kinetics from single cell transcriptomic data, we developed a Bayesian hierarchical framework to estimate the bursting kinetic parameters (i.e. transcribe rate *s*, activation rate *k*_*on*_ and deactivation rate *k*_*off*_). In the spirit of the conventional Poisson-Beta model (Vu et al., 2016), our method considered the transcriptional bursting process characterized by burst size *s*/*k*_*off*_ and burst frequency *k*_*on*_. A major limitation of Poisson-Beta model is that it neglects the remarkable noisy nature of sequencing technology^81-83^, which usually leads to overestimation of the dispersion and consequently jeopardize the validity to infer bursting kinetics. To address this issue, we reconstructed the mean-variance relationship by utilizing the Generalized additive model (GAM) that can resist and down-weight the effect of a small number of atypically high dispersion estimates especially for genes with few counts. The re-estimated dispersion *BCV* based on the re-fitted mean-variance function was then incorporated into the framework, assuming that the sampling distribution of the underlying mean *λ* is gamma. Specifically, let *y* ∼ *Poisson*(λ) denotes the observed number of mRNA molecules,

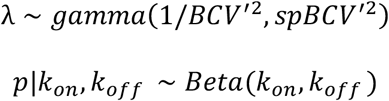

Where p denotes the fraction of time that a gene spends in the active state. With the proper choices of the priors and hyperpriors, the posterior parameters were estimated via Markov Chain Monte Carlo (MCMC) algorithm implemented in Rstan^84^.

We also formulated a differential bursting analysis framework that can further explored the change in bursting kinetics between different study conditions. After applying the MCMC method, the MCMC samples provided various representative combinations of bursting frequency and size values, while the credible differences of bursting frequency and size could be examined by computing and summarizing the changes at each combination of parameters values. Those credible differences could then be used to assess the credibility of “null value”^85^.

We applied the proposed method to investigate the differences of bursting kinetics in ANXA1^+^ DCs in COVID-19 lung compared to pDCs and cDC2s separately. With the processed scRNA-seq count data generated from previous pipeline, changes in bursting frequency and size estimates (in log_2_ scale) were computed.

### Transcriptome-based phenome-wide association study

We utilized public resources to search for CD209 related traits/disease candidate. PhenoScanncer^86^ was used to obtain associated phenotypes from genome-wide association study (GWAS) studies. Also, PhenomeXcan^87^ underlying phenome-wide association study (PheWAS) analytic framework was used to infer associations of CD209 lung expression and multiple traits, by integrating GWAS summary statistics of 4,091 traits and Genotype-Tissue Expression data from GTEx^88^ lung tissue version 8.

### Mendelian Randomization analysis

Mendelian Randomization (MR) analysis provides evidence for a causal relationship between exposure and outcome. To infer the causal effect of plasma expression of ACE2 and DC-SIGN on COVID-19 risk and severity, we applied the TwoSampleMR R package^89^ for MR analysis. For exposures of protein expression of SARS-CoV-2 receptors, we obtained publicly available plasma protein QTL summary statistics from the study of Sun *et al*.^45^. For outcomes of COVID-19 risk and severity, we separately required GWAS summary statistics from GRASP (https://grasp.nhlbi.nih.gov/COVID-19GWASResults.aspx; files with name “ALLwhites” at 06-05-2020) for COVID-19 risk, and that from the study of Ellinghaus et al.^46^ for COVID-19 severity. Independent genetic instruments of exposures were selected for MR analysis based on strict criteria of minor allele frequency (MAF) > 0.05, linkage disequilibrium (LD) r^2^ < 0.01 within a clumping window of 10,000 kb, and beyond genome-wide significance (P < 5×10^−8^). As there were no genetic instruments of plasma ACE2, the significance threshold for selecting instruments was set as P < 10^−4^. Ultimately, we found 3, 60, and 1 SNPs as independent genetic instruments for plasma CD209, ACE2, and CLEC4M, respectively. Inverse-variance-weighted (IVW), weighted median, and MR-Egger regression methods were used to calculate effect size (*β*) and corresponding standard error (SE). The Wald ratio method was used if only one genetic instrument remained. Heterogeneity was estimated using MR-Egger and IVW methods.

Directional pleiotropy was estimated via MR-Egger intercept test.

### Software

All data management, statistical analyses and visualizations were accomplished using R software version 4.0.2.

## Data Availability

The data derived from MEARDS was accessible upon request to Dr. David C. Christiani Data from other studies were openly available to the public without registration or application for access. Processed single cell transcriptomics data were available for visualization and exploration in SCANNER (https://www.thecailab.com/scanner/). The hosted data include three datasets from the normal airway samples from ever-smokers and non-smokers, as well as four datasets from the DCs of nasopharynx/pharynx, BULF and PBMC samples from COVID-19 patients and healthy people.

## Data availability

The data derived from MEARDS was accessible upon request to Dr. David C. Christiani. Data from other studies were openly available to the public without registration or application for access. Processed single cell transcriptomics data were available for visualization and exploration in SCANNER^48^ (https://www.thecailab.com/scanner/). The hosted data include three datasets from the normal airway samples from ever-smokers and non-smokers, as well as four datasets from the DCs of nasopharynx/pharynx, BULF and PBMC samples from COVID-19 patients and healthy people.

## Ethical oversight

All the data were de-identified and study approval has been stated in original studies. The MEARDS study was reviewed and approved by institutional review boards of Massachusetts General Hospital (MGH) and Harvard School of Public Health (project approval number: 1999P0086071/MGH).

## Supplementary File

**Figure S1.**
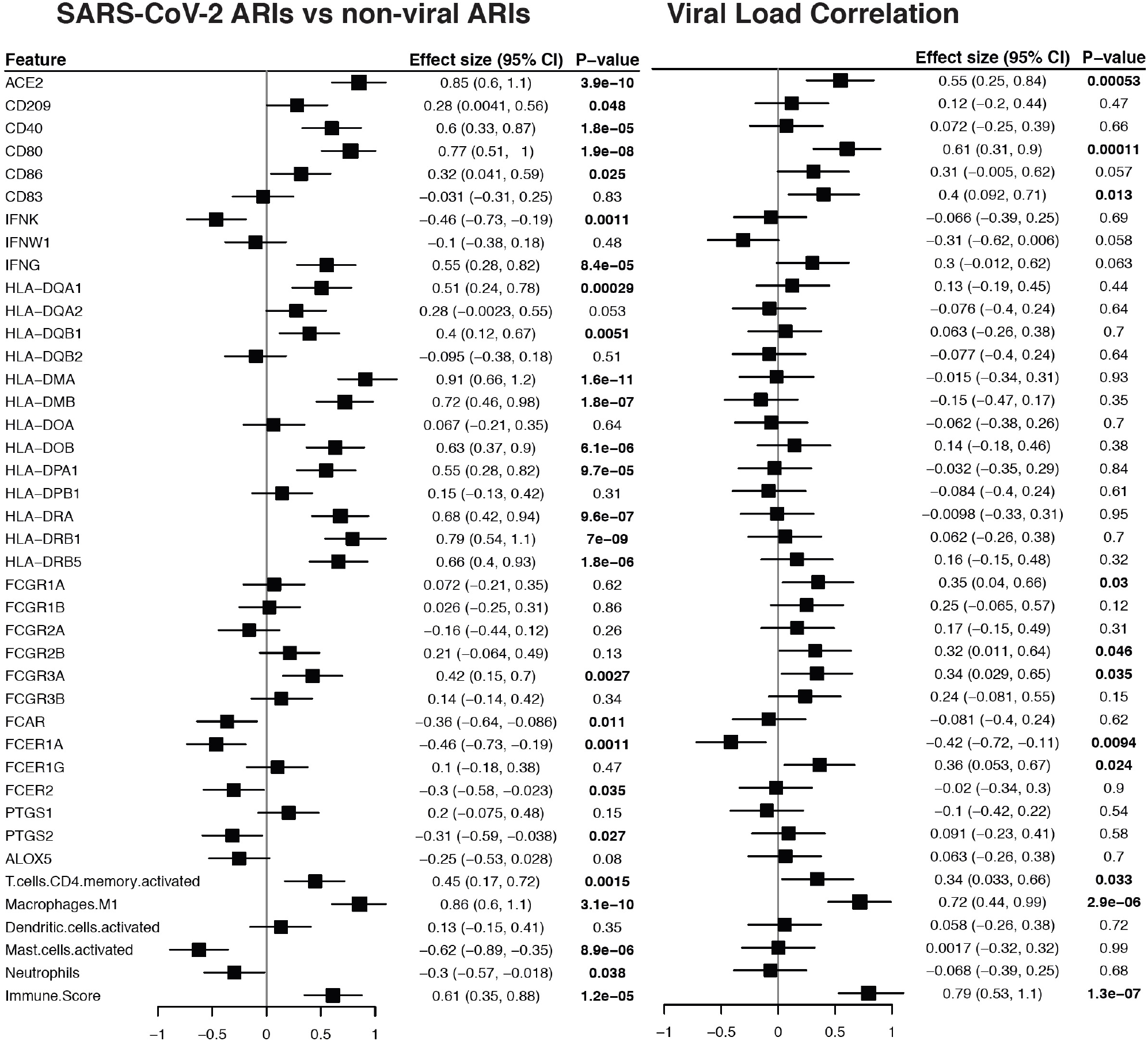
The association of SARS-CoV-2 receptors and immune factors with SARS-CoV-2 infection and viral load in nasopharynx/pharynx samples of ARIs. For each feature, results of comparisons between SARS-CoV-2 ARIs and non-viral ARIs are shown on the left and correlations with SARS-CoV-2 viral load are shown on the right. For each feature, the estimated coefficient and 95% confidence intervals (CIs) are plotted. Significant P values which are less than 0.05 were shown in bold.

**Figure S2.**
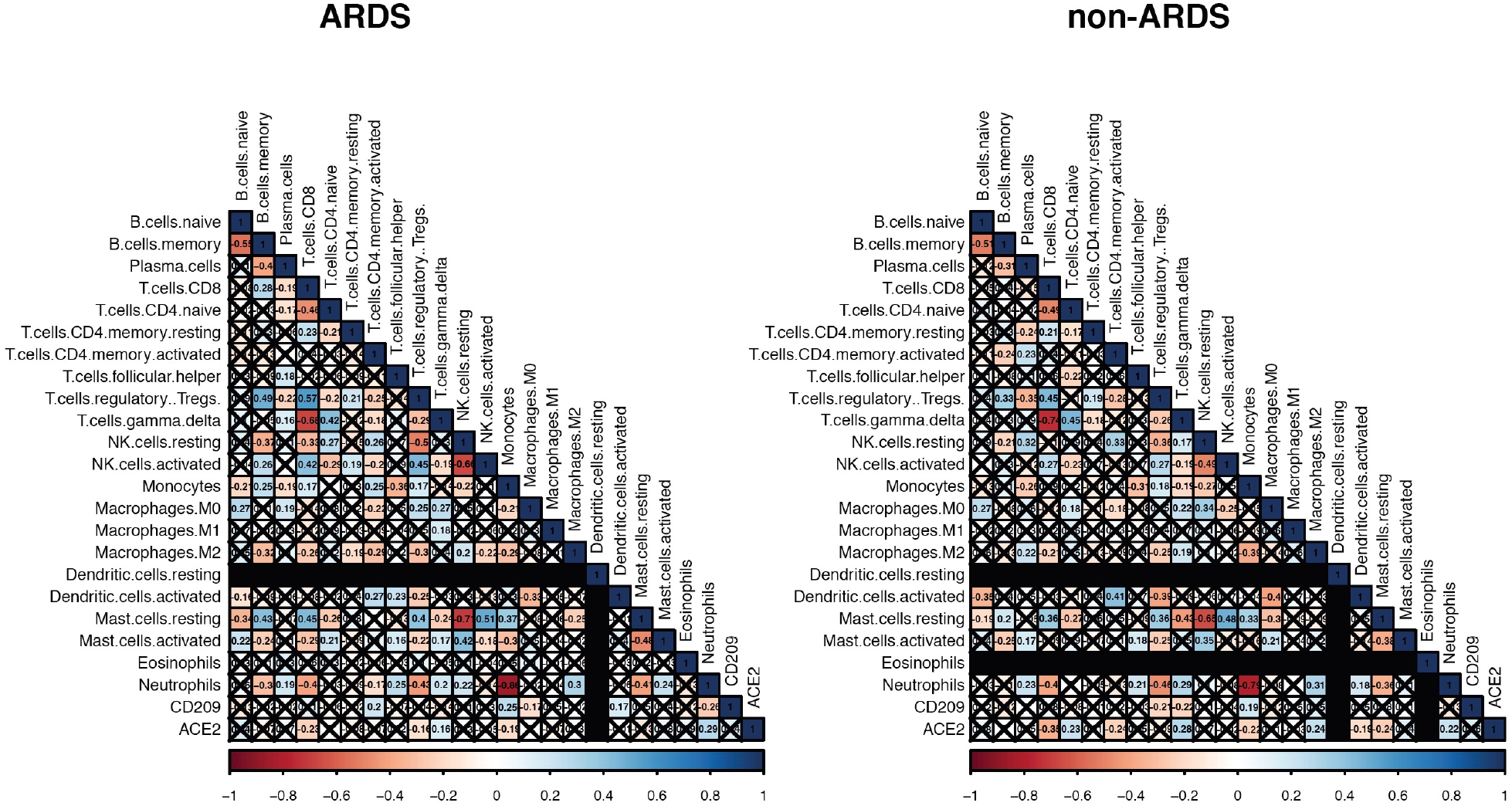
Correlation matrix among immune cell composition and the expression of *ACE2, CLEC4M* and *CD209* in ARDS and non-ARDS blood samples. Pairwise similarity (Spearman correlation) among the expression of candidate genes and 22 immune cells proportions in ARDS blood samples is shown on the left, and that in non-ARDS blood samples is shown on the right.

**Figure S3.**
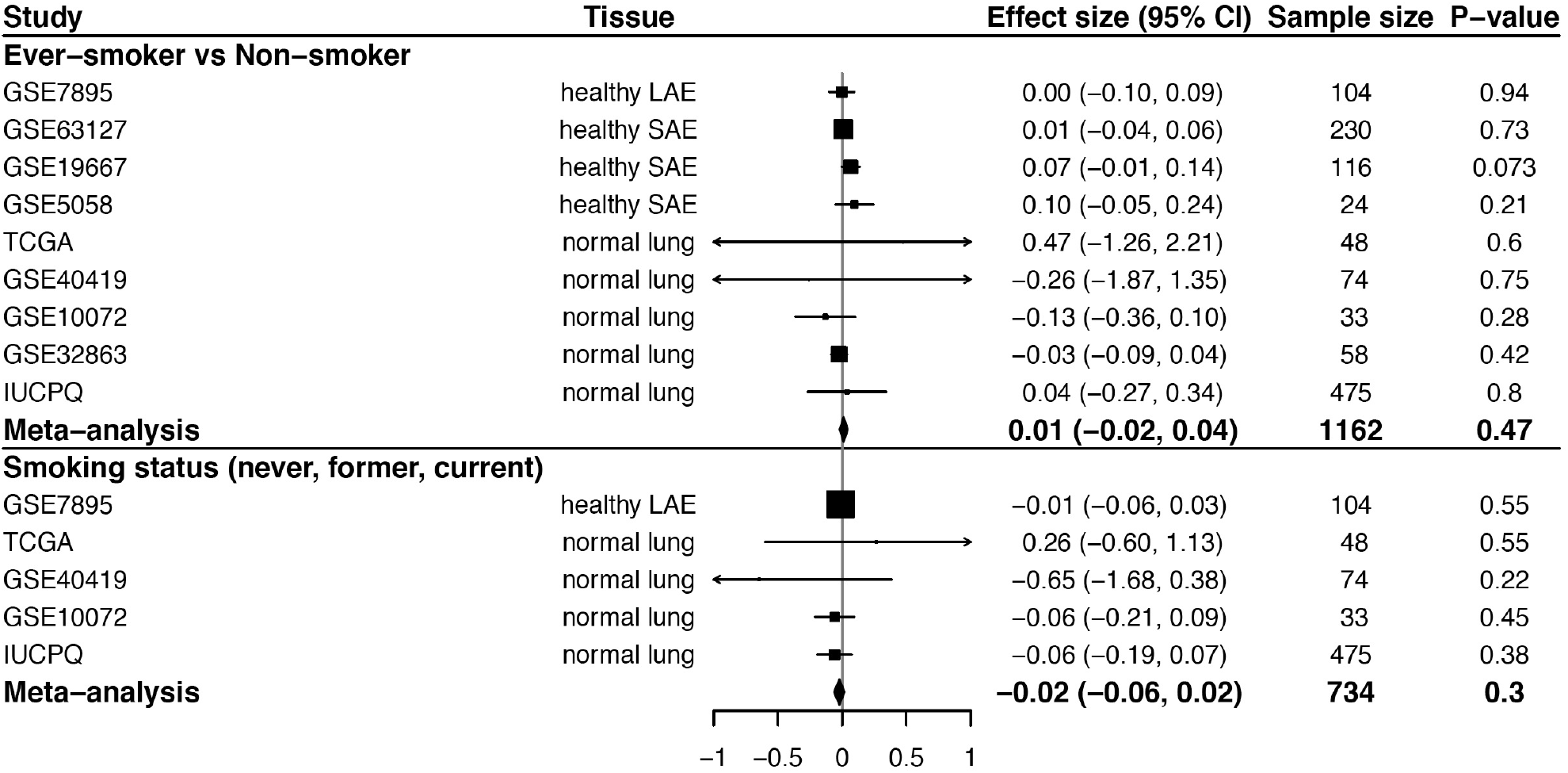
The forest plots for the effects of smoking on *ACE2* pulmonary gene expression. For each cell type, the top panel shows a comparison of ever-smoker (including current and former smokers) and nonsmoker groups, and the bottom panel shows the association of the gene expression with smoking status (never, former, or current smoker). For each study, the estimated effect size and 95% confidence intervals (CIs) are plotted. The size of the squares is proportional to the weights, which were estimated by the standard “inverse-variance” method for random-effects models in meta-analysis.

**Figure S4.**
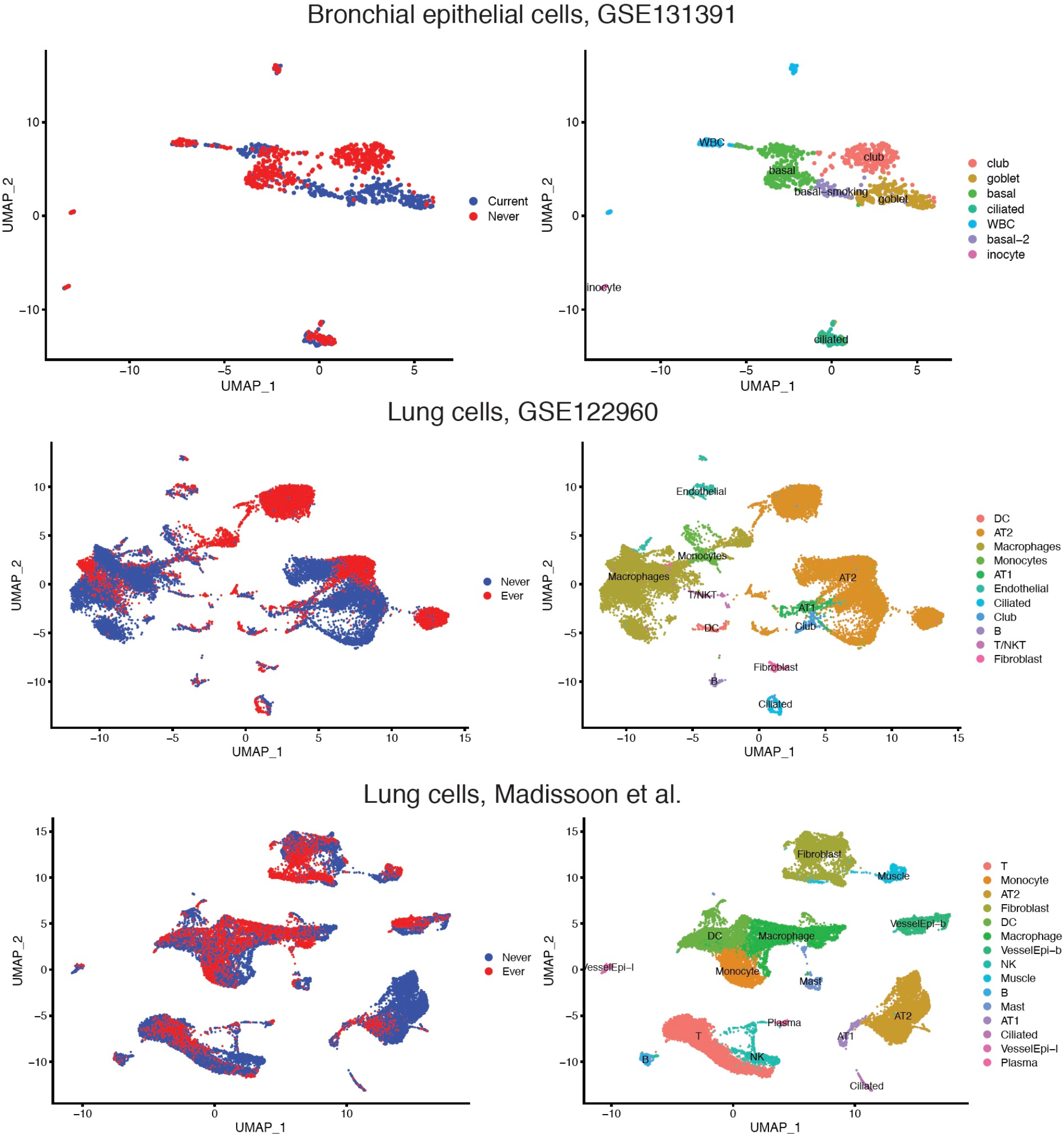
Clusters of single-cell transcriptomics of bronchial epithelium and lung cells. For each dataset, the left shows UMAP embedding of single-cell transcriptome profiles from never smokers and current or ever smokers. The right shows identified cell types.

**Figure S5.**
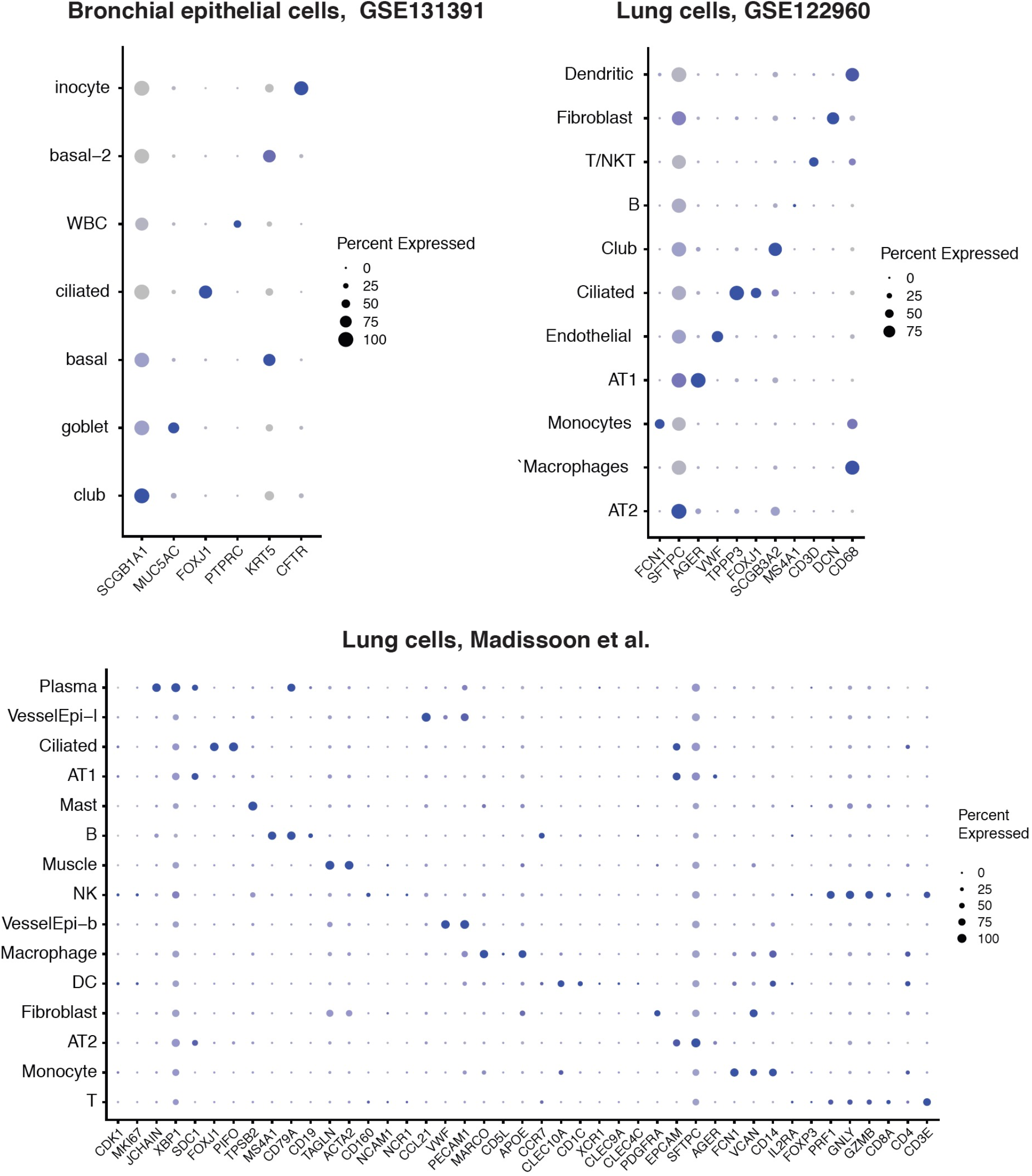
Expression profiles of cell type markers in scRNA-seq data of bronchial epithelium and lung cells. For each marker, detection rate (showed by dot size, larger dot indicates higher detection rate) and expression level (showed by dot color which is status specific, darker indicates higher expression level) are shown in identified cell types.

**Figure S6.**
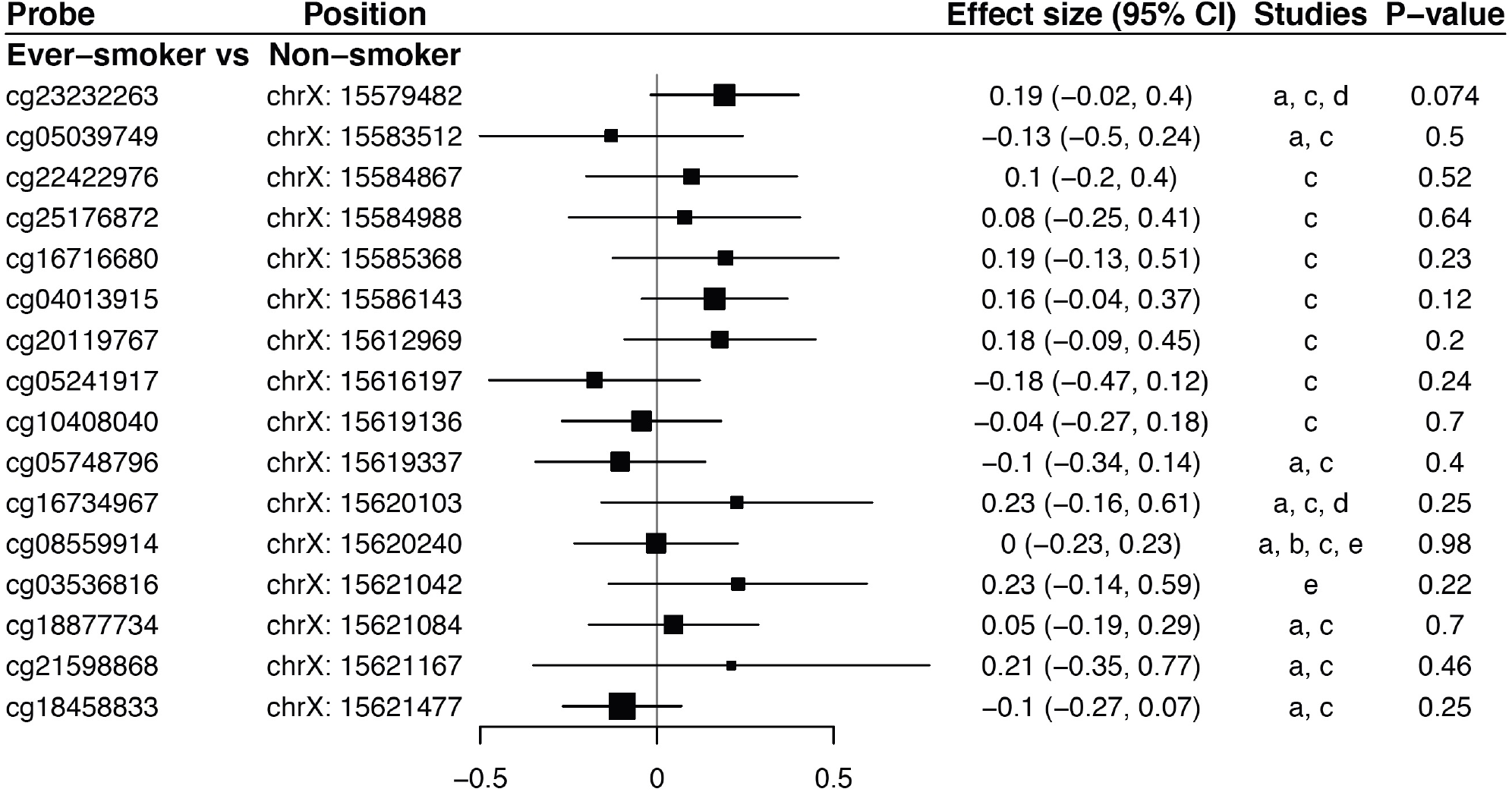
The forest plot for the effects of smoking on *ACE2* related methylation probes in normal lung tissues. For each probe, pooled effect size, 95% CI and P values were calculated by meta-analysis from available studies, including a: TCGA, b: GSE32861, c: GSE133062, d: GSE92511 and e: GSE139032. For each study, the estimated effect size and 95% confidence intervals (CIs) are plotted. The size of the squares is proportional to the weights, which were estimated by the standard “inverse-variance” method for random-effects models in meta-analysis.

**Figure S7.**
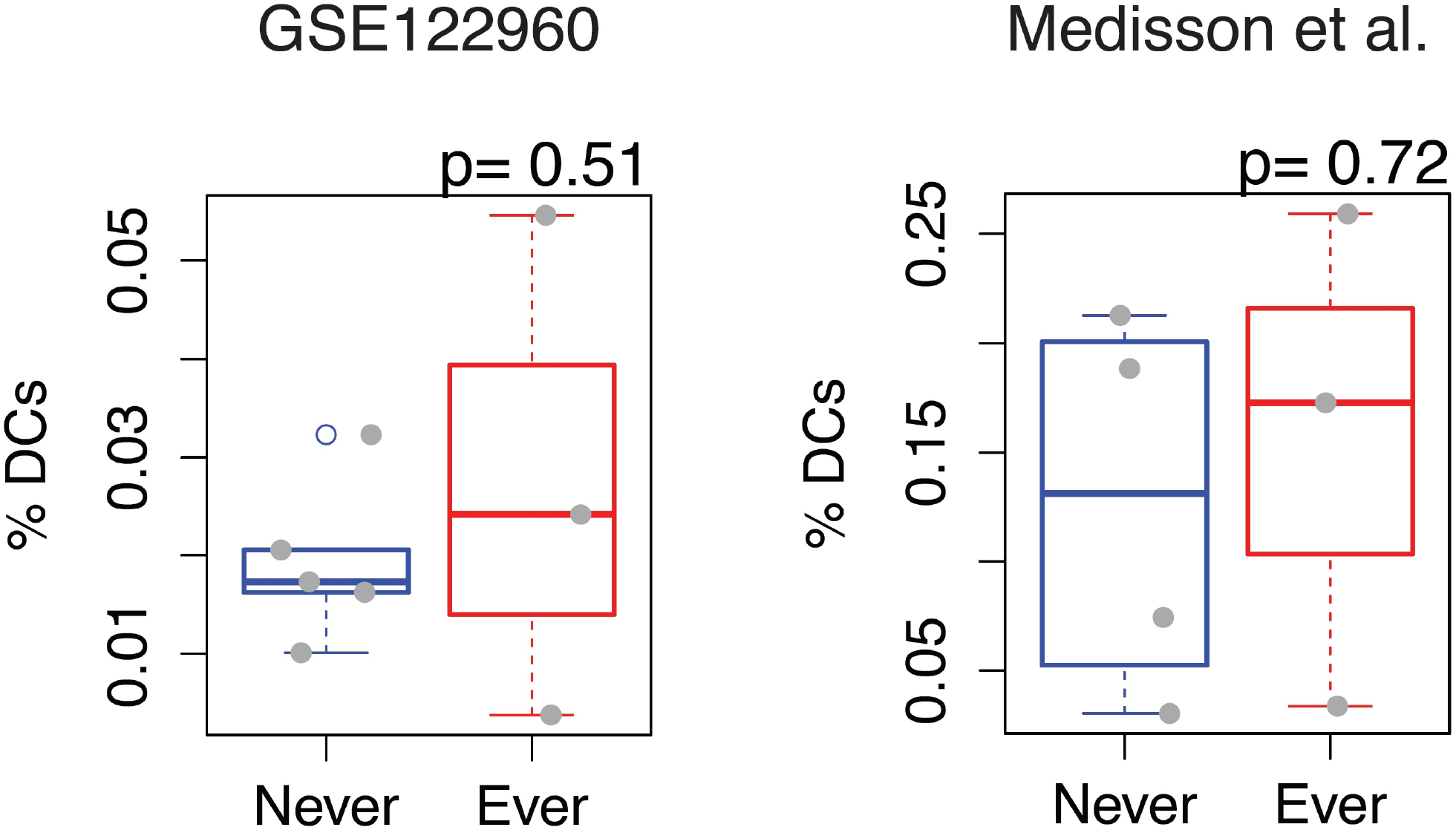
The proportion of DCs in WBC of lungs of never and ever smokers. The proportion of DCs were enumerated from the scRNA-seq data of lung samples of each participant recruited in two independent studies.

**Figure S8.**
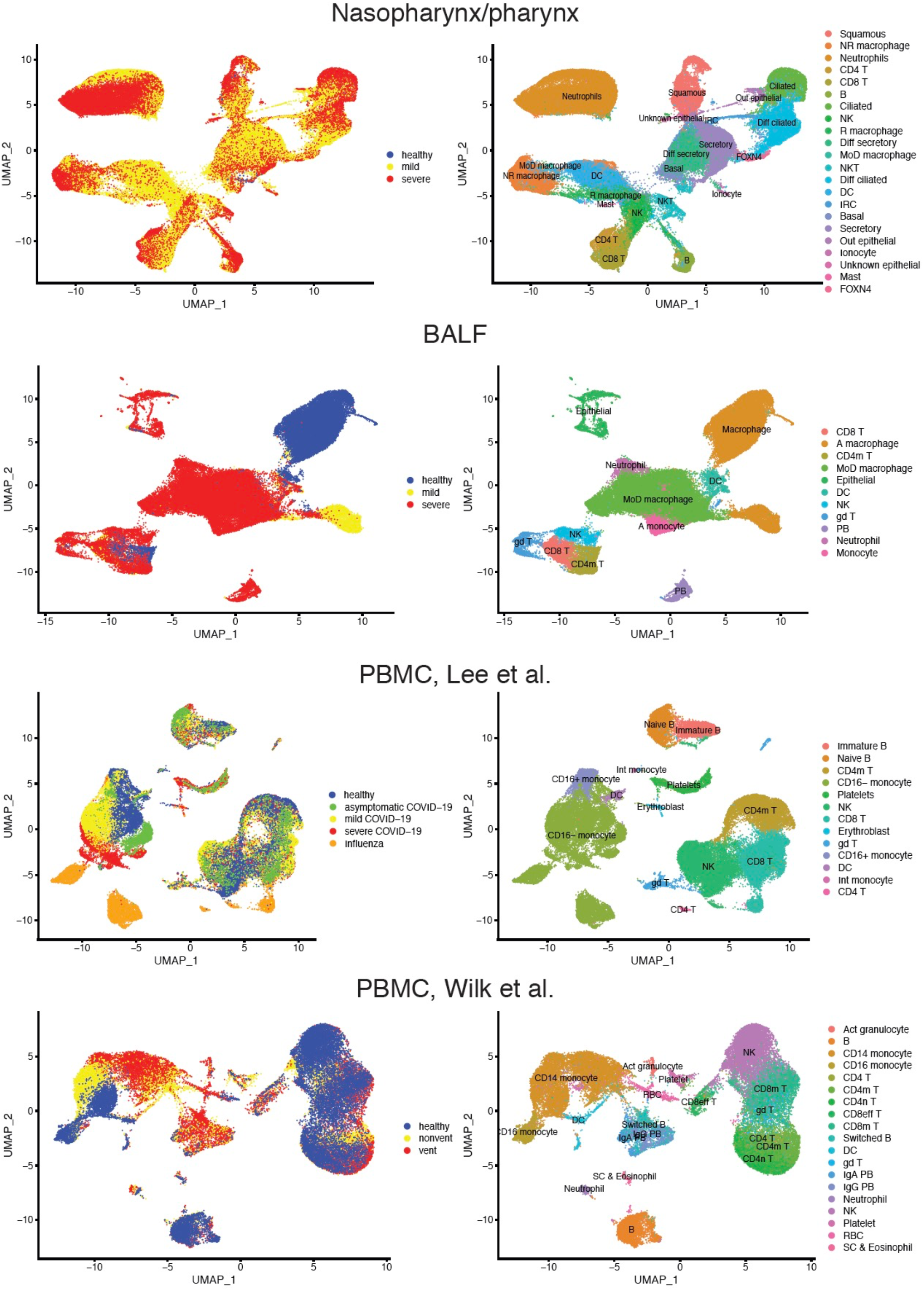
Clusters of single-cell transcriptomics of nasopharynx/pharynx, BALF and PMBC samples from COVID-19 cases and healthy controls. For each dataset, the left shows UMAP embedding of single-cell transcriptome profiles from COVID-19 mild and severe cases, and healthy controls. The right shows identified cell types.

**Figure S9.**
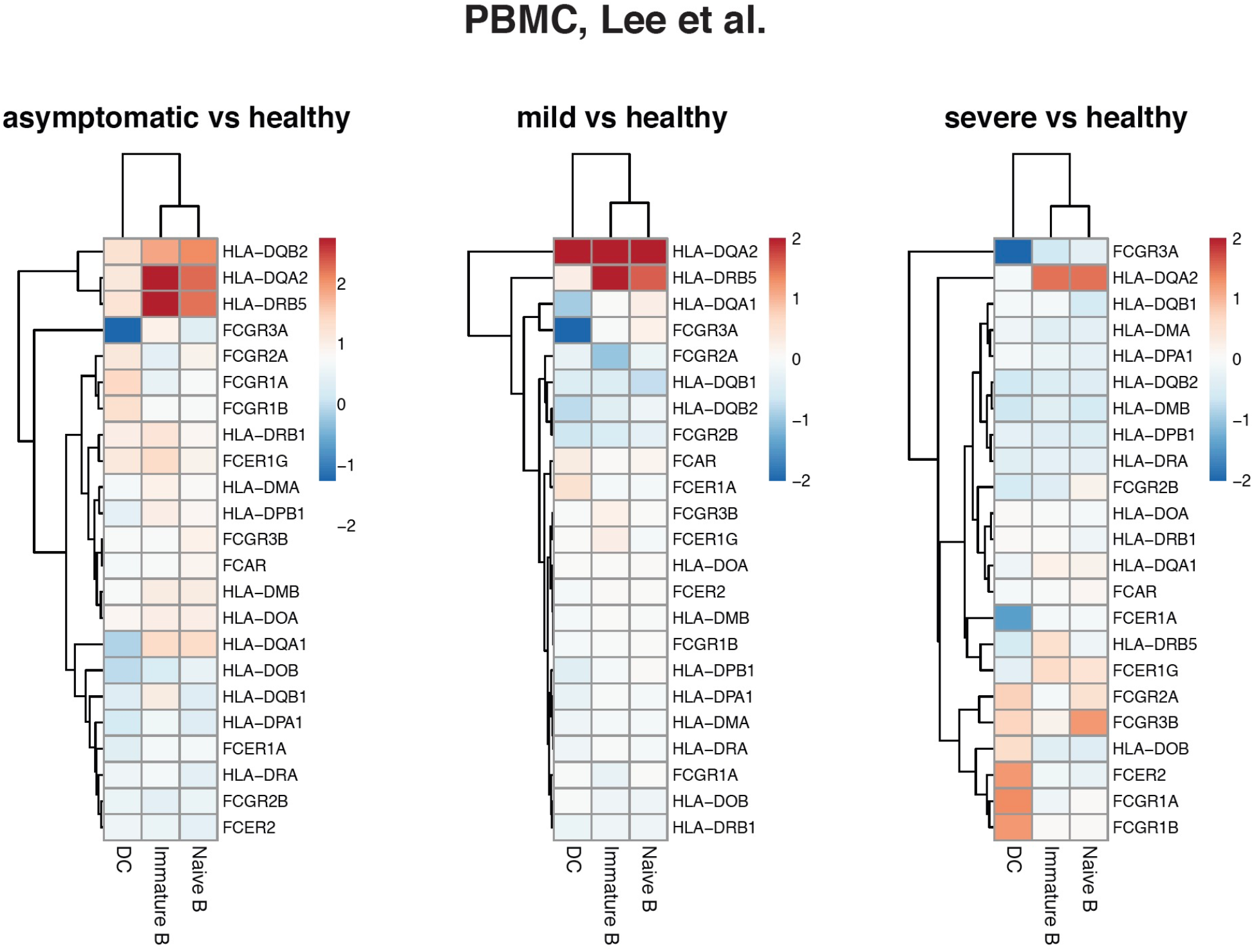
The dysregulation of gene expression of MHCII and FcRs in APCs in blood with COVID-19. For each gene, the alteration of gene expression in asymptomatic, mild and severe COVID-19 cases compared to healthy controls are visualized in heatmaps. Red indicates upregulation and blue indicates downregulation. The scale corresponds to log_2_FC. The PBMC scRNA-seq dataset of Lee et al. was analyzed.

**Figure S10.**
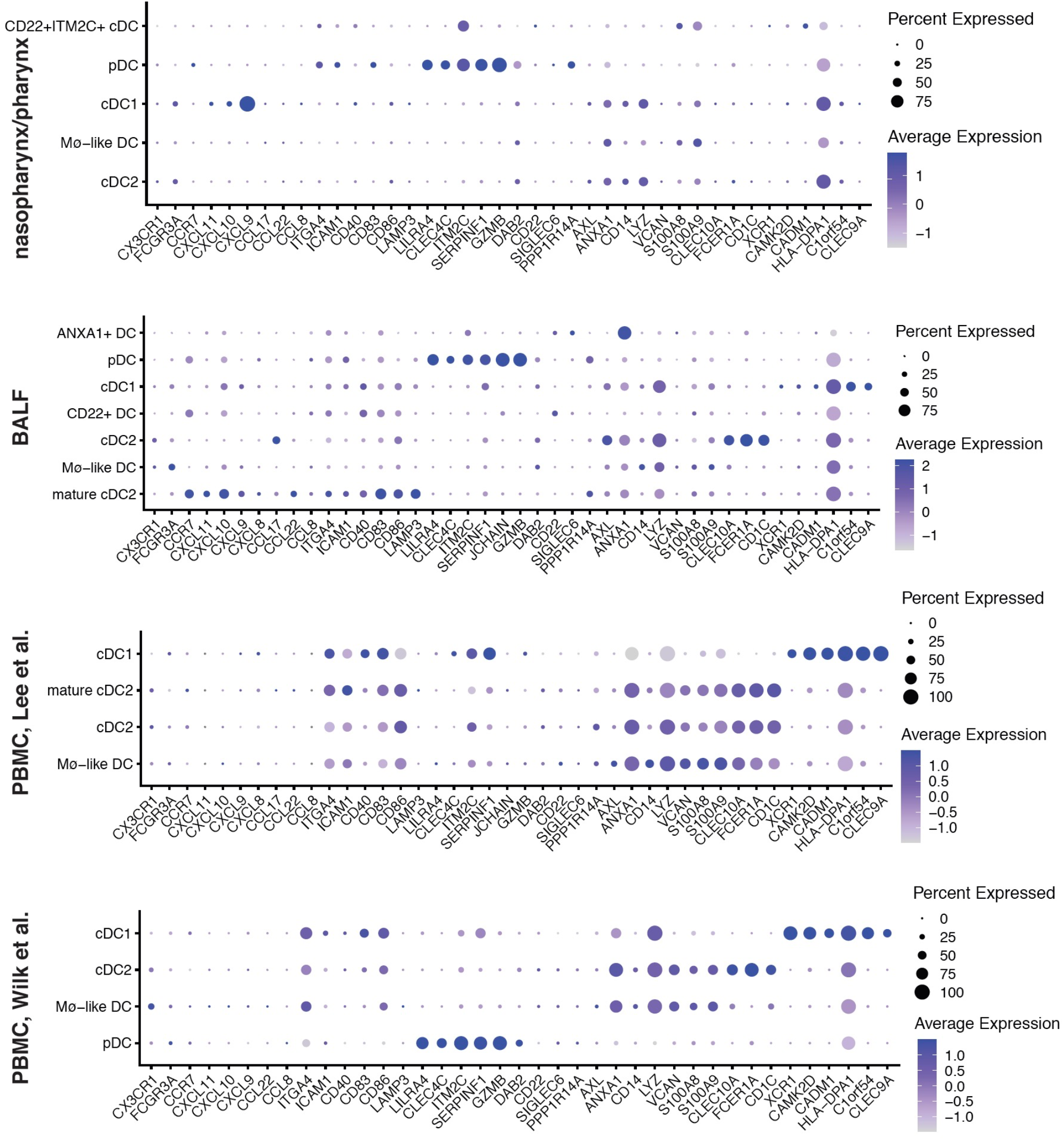
Expression profiles of DC subset markers in four scRNA-seq datasets of nasopharynx/pharynx, BALF and PBMC samples from COVID-19 patients and healthy controls. For each dataset, the detection rate (showed by dot size, larger dot indicates higher detection rate) and expression level (showed by dot color, darker indicates higher expression level) of each cell-type marker are shown in identified DC subtypes.

**Figure S11.**
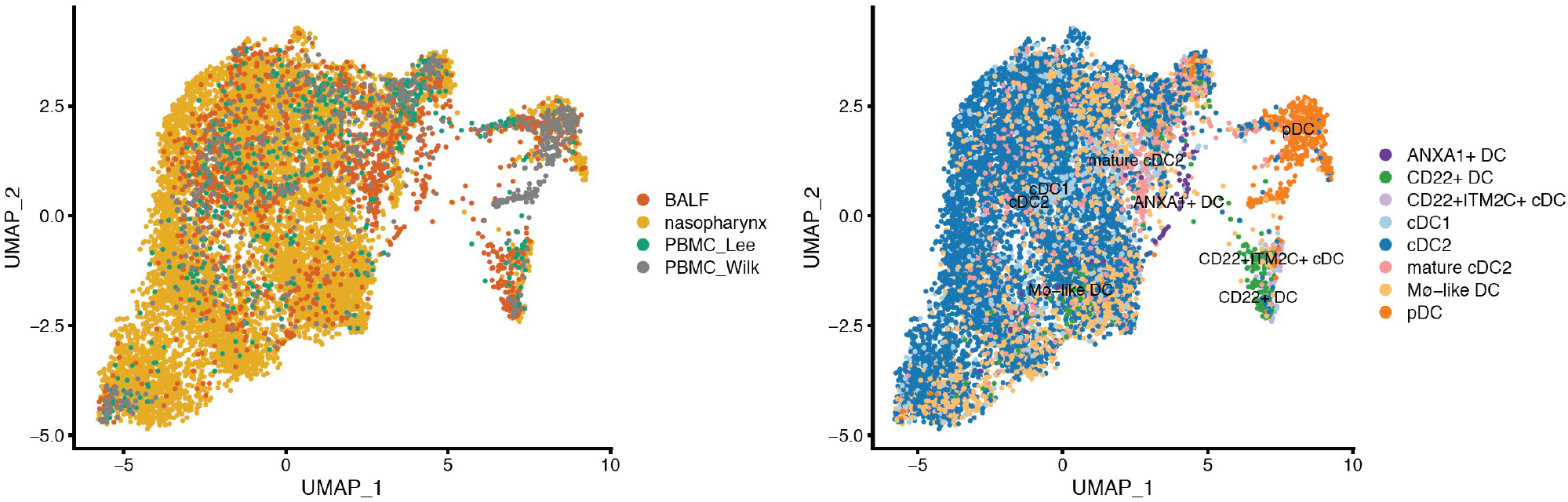
Integrated single-cell transcriptomics of nasopharynx/pharynx, BALF and PMBC samples from COVID-19 cases and healthy controls. The left shows UMAP embedding of single-cell transcriptome profiles from COVID-19 mild and severe cases, and healthy controls. The right shows identified cell types.

**Figure S12.**
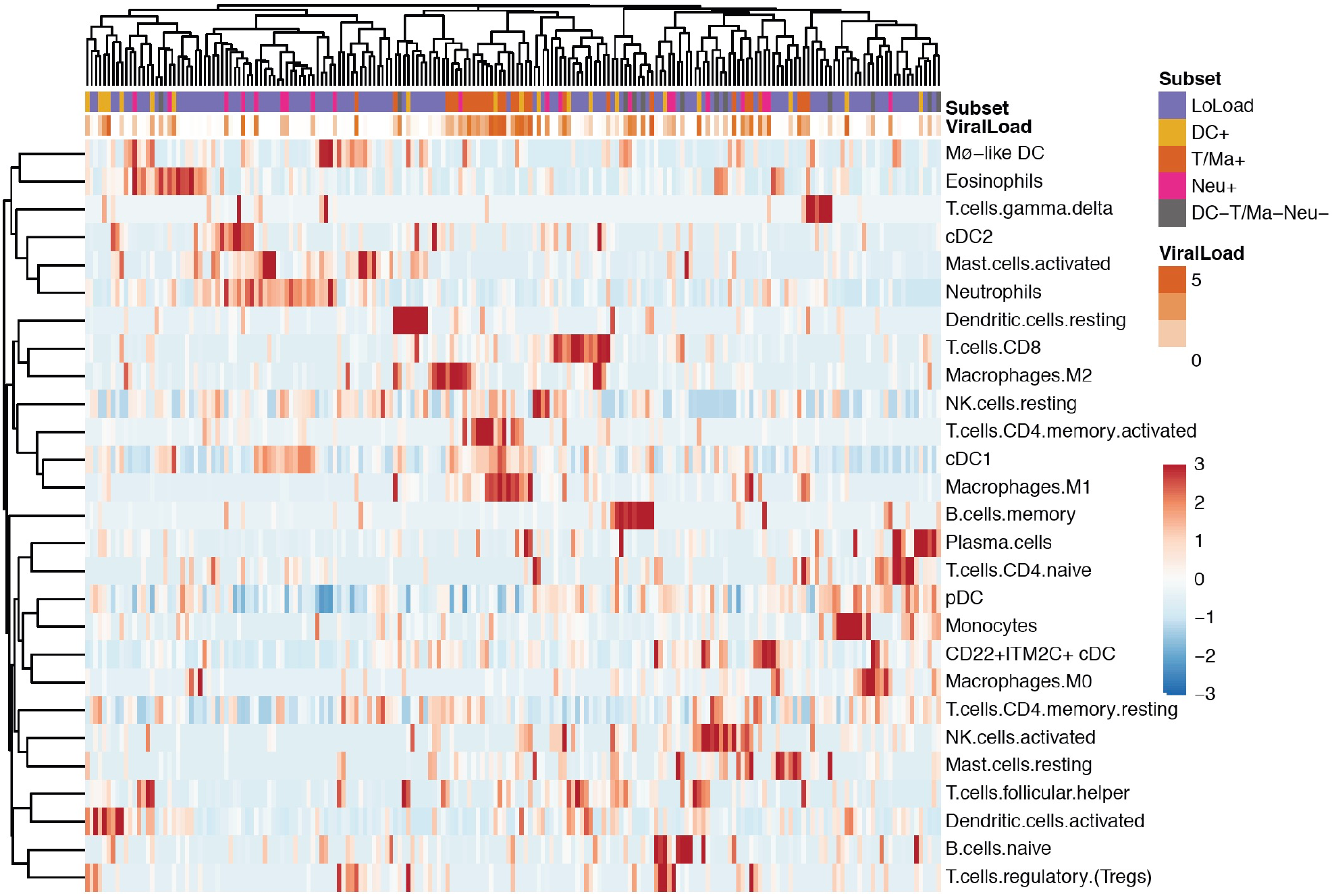
Hierarchical clustering and heatmap of immune cell composition in ARI nasopharynx/pharynx samples from COVID-19 cases and healthy controls. For each cell type, red indicates relatively higher proportion and blue indicates relatively higher proportion across samples.

**Figure S13.**
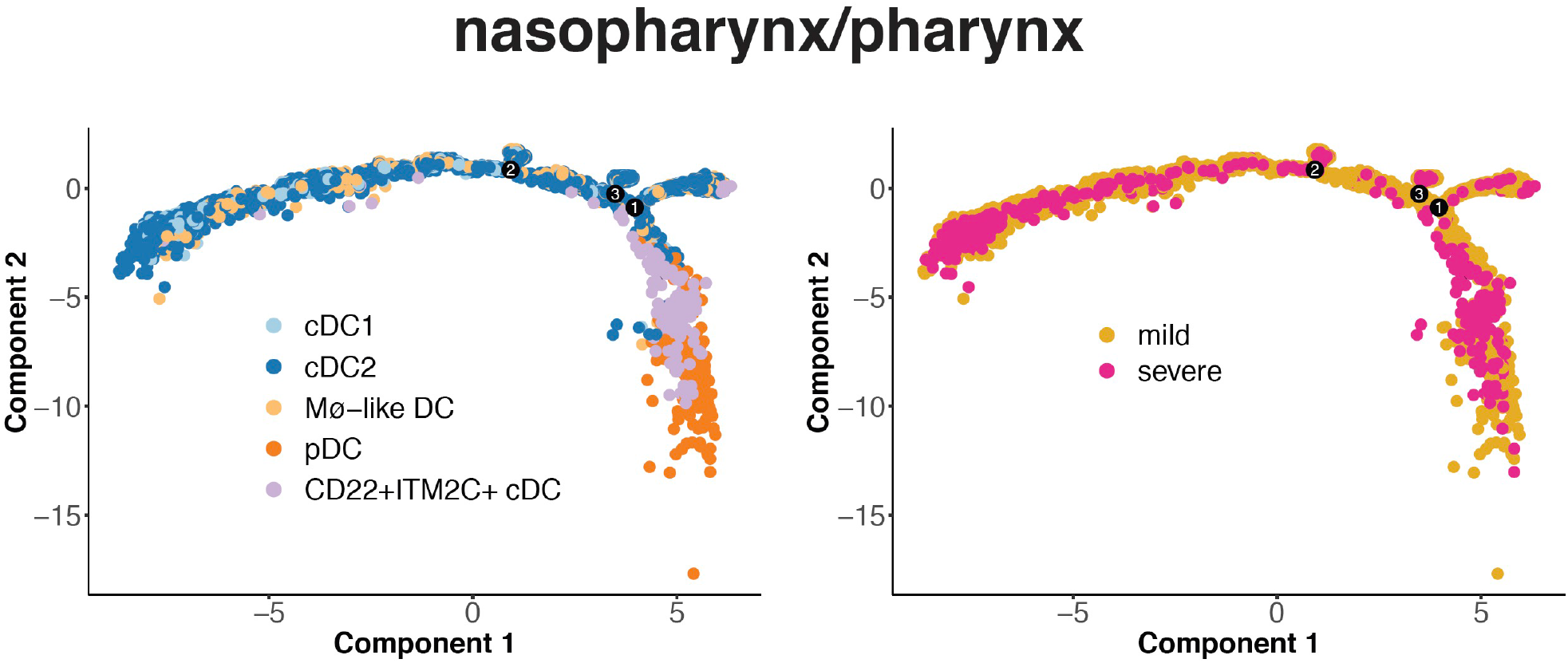
Single-cell trajectory of DCs in COVID-19 nasopharynx/pharynx samples. The right shows ordered cells from COVID-19 mild and severe cases. The left shows ordered cells labeled with identified cell types.

**Figure S14.**
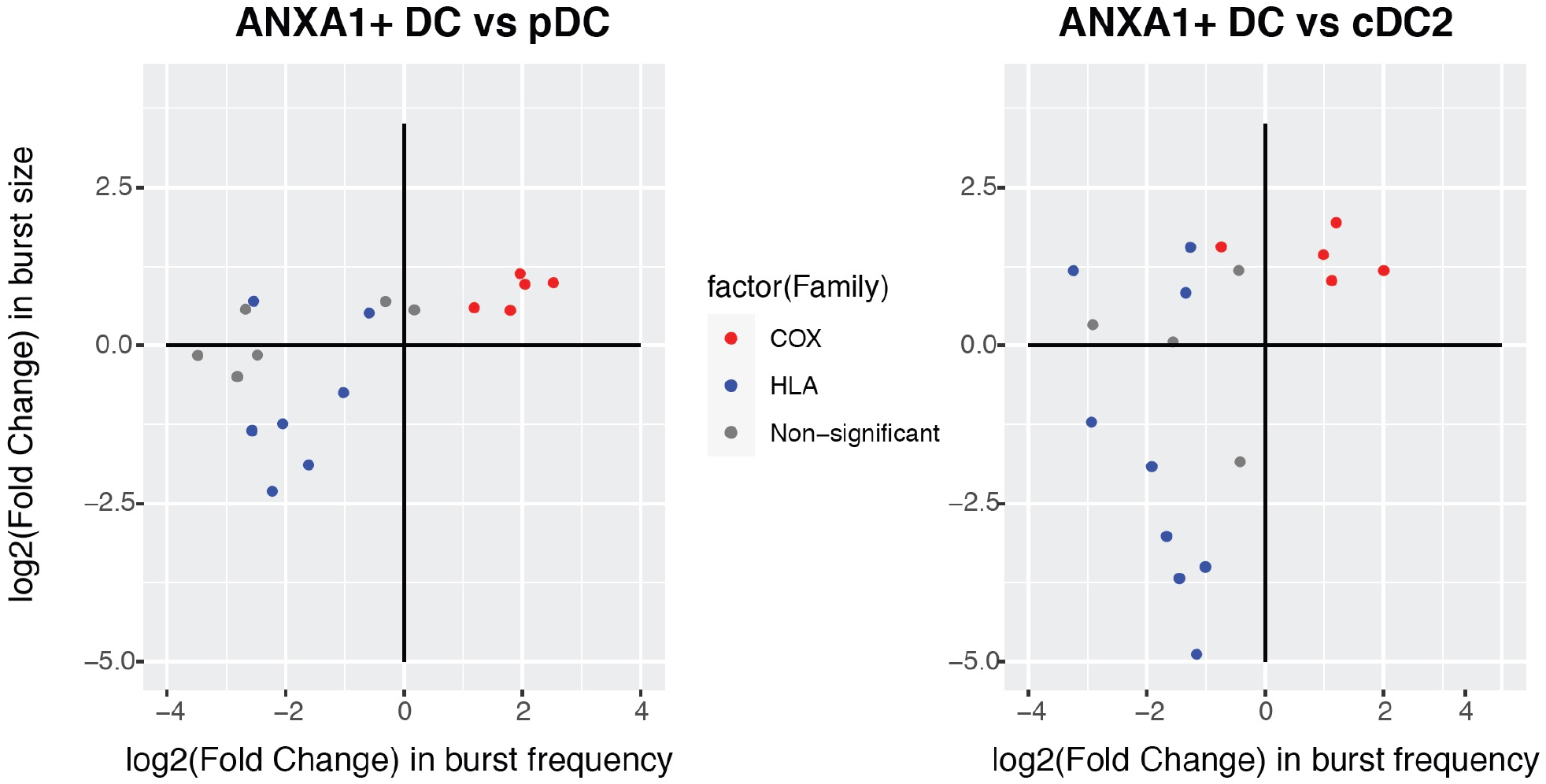
Transcriptional bursting profiles in different DC subsets. The changes in burst frequency and burst size of HLA genes (blue) and Cox genes (red) in ANXA1+DC compared to pDC (left) and cDC2 (right) are plotted in log_2_ scale.

**Figure S15.**
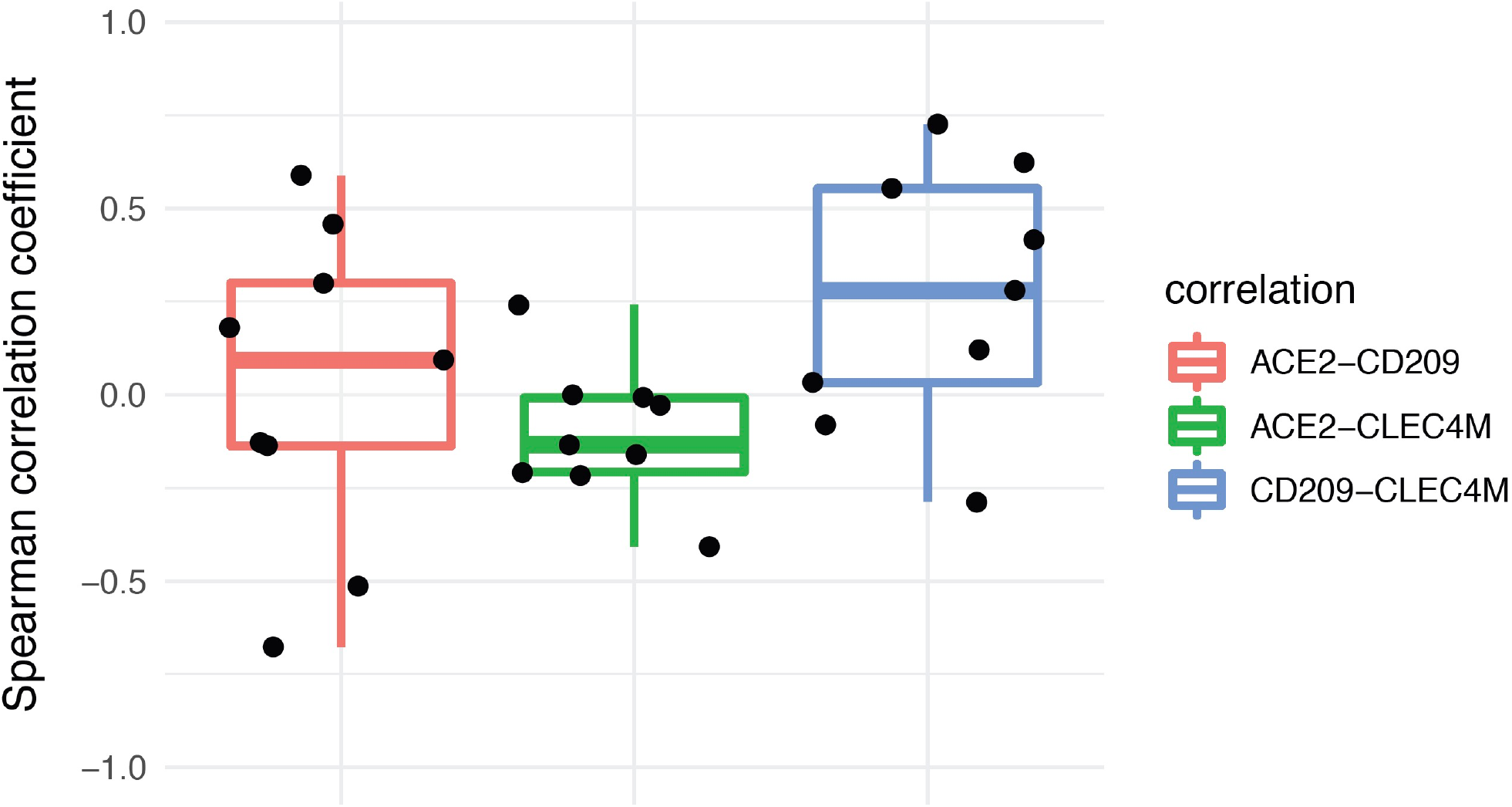
Correlation of genes *ACE2, CLEC4M* and *CD209* in normal lung tissues. Spearman correlation coefficients for expression of each pair of genes were shown in boxplot.

**Table S1.**
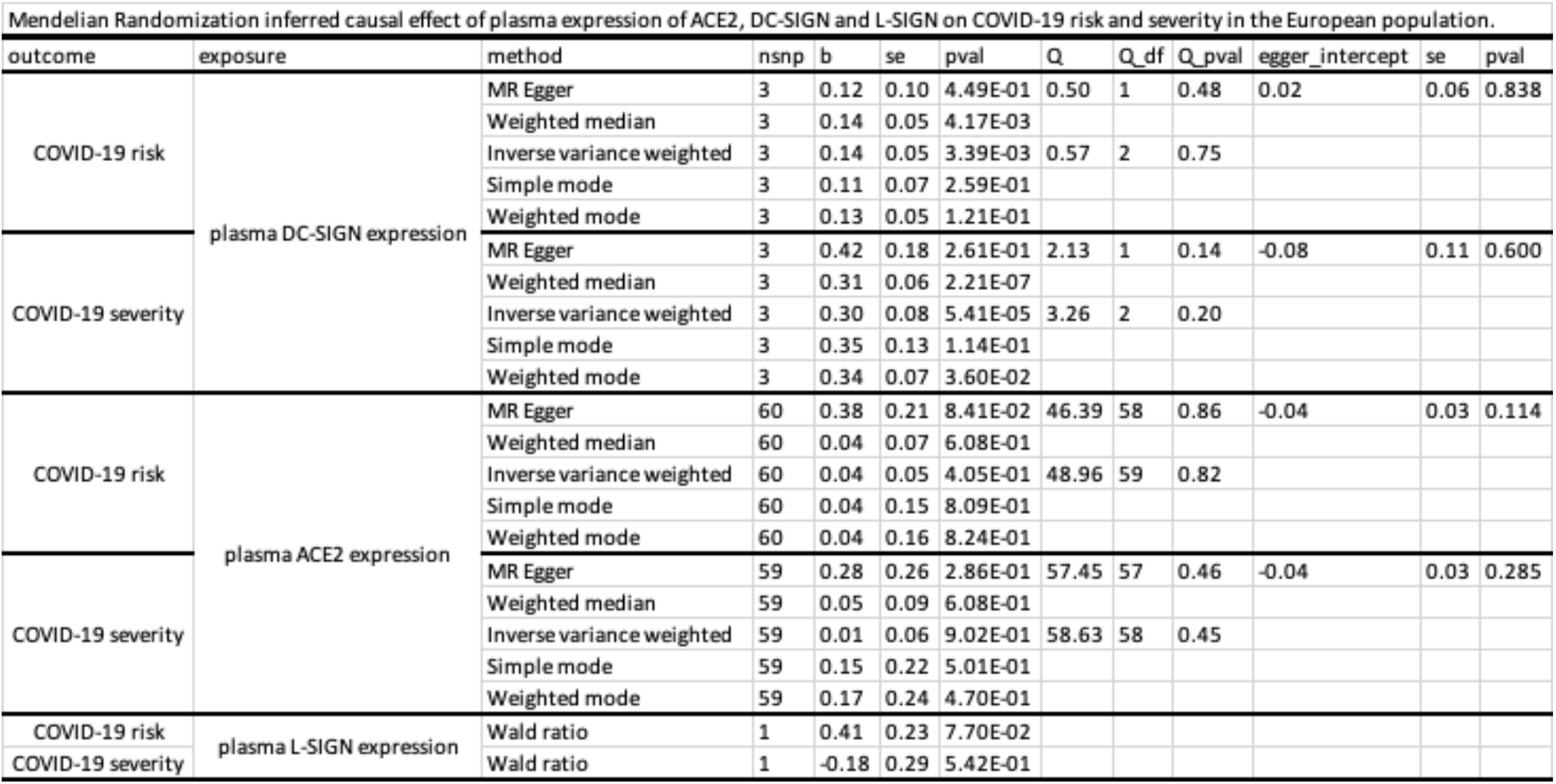
Mendelian Randomization inferred causal effect of plasma expression of ACE2, DC-SIGN and L-SIGN on COVID-19 risk and severity in the European population.

**Table S2.**
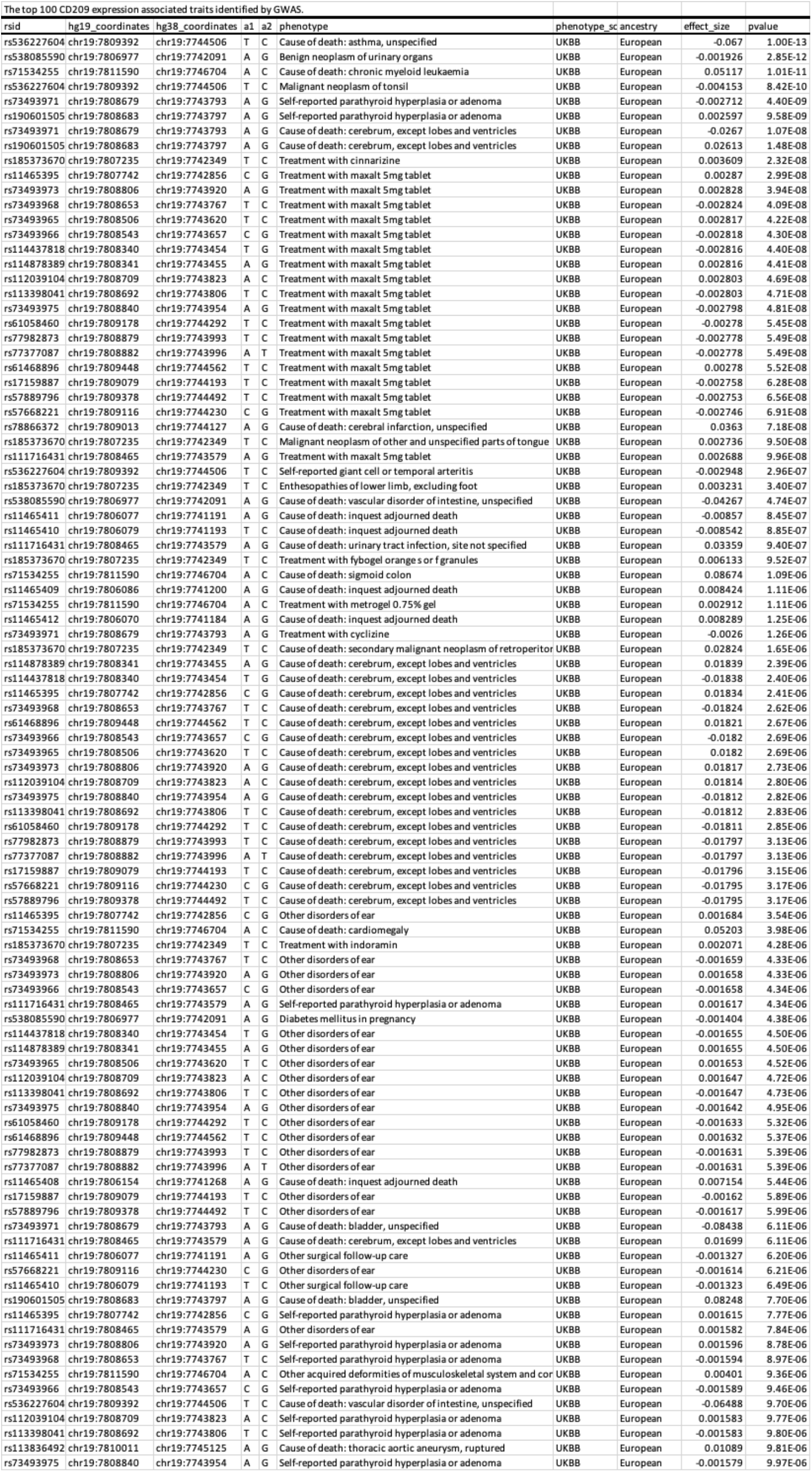
The top 100 CD209 expression associated traits identified by GWAS.

**Table S3.**
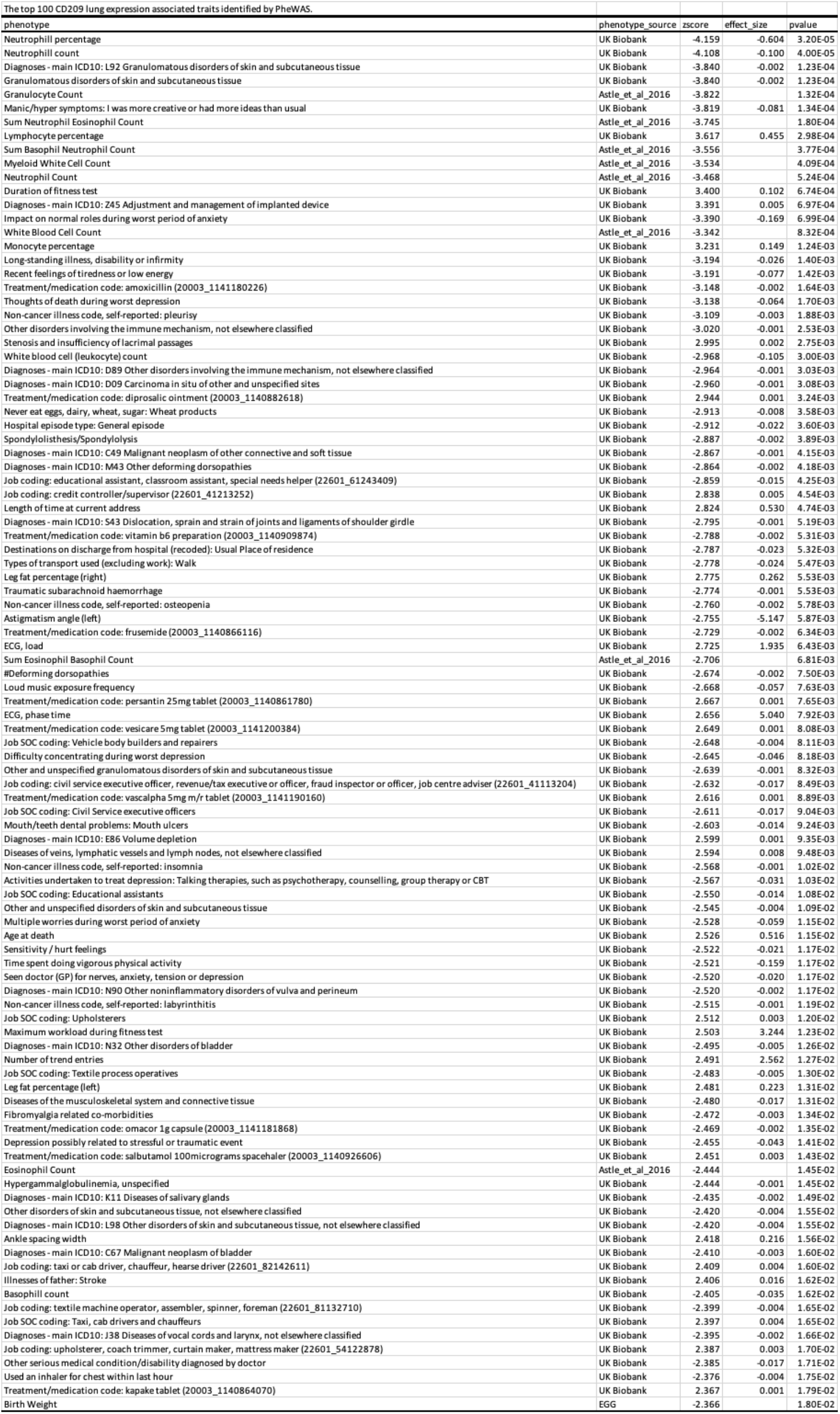
The top 100 CD209 lung expression associated traits identified by PheWAS.

**Table S4.** Mendelian Randomization inferred causal effect of plasma expression of DC-SIGN on cell count and percentage.

|  |  |  |  |  |  |  |
| --- | --- | --- | --- | --- | --- | --- |
| outcome | exposure | method | nsnp | b | se | pval |
| White blood cell (leukocyte) count \|\| id:ukb-d-30000_irnt | plasma DC-SIGN expression | Inverse variance weighted | 3 | -0.0305717 | 0.00284163 | 5.40E-27 |
| Neutrophil count \|\| id:ukb-d-30140_irnt | plasma DC-SIGN expression | Inverse variance weighted | 3 | -0.0274776 | 0.00332404 | 1.38E-16 |
| Red blood cell count \|\| id:ieu-a-275 | plasma DC-SIGN expression | Inverse variance weighted | 3 | -0.0140248 | 0.00271122 | 2.31E-07 |
| Lymphocyte percentage \|\| id:ukb-d-30180_irnt | plasma DC-SIGN expression | Inverse variance weighted | 3 | 0.01685935 | 0.00417076 | 5.29E-05 |
| Granulocyte count \|\| id:ebi-a-GCST004614 | plasma DC-SIGN expression | Inverse variance weighted | 2 | -0.0422213 | 0.01085916 | 0.00010104 |
| Sum neutrophil eosinophil counts \|\| id:ebi-a-GCST004613 | plasma DC-SIGN expression | Inverse variance weighted | 2 | -0.0412766 | 0.01084251 | 0.00014071 |
| Myeloid white cell count \|\| id:ebi-a-GCST004626 | plasma DC-SIGN expression | Inverse variance weighted | 2 | -0.0397515 | 0.01088796 | 0.00026126 |
| Sum basophil neutrophil counts \|\| id:ebi-a-GCST004620 | plasma DC-SIGN expression | Inverse variance weighted | 2 | -0.0389864 | 0.01085165 | 0.00032731 |
| Granulocyte percentage of myeloid white cells \|\| id:ebi-a-GCST004608 | plasma DC-SIGN expression | Inverse variance weighted | 2 | -0.0385173 | 0.01080265 | 0.0003631 |
| Neutrophil count \|\| id:ebi-a-GCST004629 | plasma DC-SIGN expression | Inverse variance weighted | 2 | -0.03781 | 0.01083361 | 0.00048291 |
| White blood cell count \|\| id:ebi-a-GCST004610 | plasma DC-SIGN expression | Inverse variance weighted | 2 | -0.0369582 | 0.01084099 | 0.00065174 |
| Lymphocyte count \|\| id:ukb-d-30120_irnt | plasma DC-SIGN expression | Inverse variance weighted | 3 | -0.0109447 | 0.00348627 | 0.0016931 |
| Red blood cell (erythrocyte) count \|\| id:ukb-d-30010_irnt | plasma DC-SIGN expression | Inverse variance weighted | 3 | -0.0304825 | 0.00983767 | 0.00194473 |
| Monocyte percentage of white cells \|\| id:ebi-a-GCST004609 | plasma DC-SIGN expression | Inverse variance weighted | 2 | 0.03090055 | 0.01174645 | 0.00852271 |
| Eosinophil counts \|\| id:ebi-a-GCST004606 | plasma DC-SIGN expression | Inverse variance weighted | 2 | -0.0280988 | 0.01078562 | 0.0091817 |
| Monocyte count \|\| id:ukb-d-30130_irnt | plasma DC-SIGN expression | Inverse variance weighted | 3 | -0.0422452 | 0.01664811 | 0.01116349 |
| Neutrophil percentage of white cells \|\| id:ebi-a-GCST004633 | plasma DC-SIGN expression | Inverse variance weighted | 2 | -0.024563 | 0.01079162 | 0.02283885 |
| Eosinophil percentage \|\| id:ukb-d-30210_irnt | plasma DC-SIGN expression | Inverse variance weighted | 3 | -0.0092451 | 0.00411516 | 0.0246658 |
| Lymphocyte percentage of white cells \|\| id:ebi-a-GCST004632 | plasma DC-SIGN expression | Inverse variance weighted | 2 | 0.02273265 | 0.01077233 | 0.03483415 |

## Acknowledgments

This work was supported by University of South Carolina COVID-19 research initiative (G.C.).

## Author Contributions

Conceptualization, G.C. and F.X.; Methodology, G.C. and F.X.; Software/Data Analysis, G.C., Y.B., M.D, F.Q., X.Y. X.L, X.C.; Investigation, G.C., H.A., M.A., X.Z, J.Z., C.Y., C.C., M.N., P.N., D.C., M.W., C.A, and F.X.; Writing – Original Draft, G.C., X.Z., J.Z. and F.X.; Writing – Review & Editing, G.C., Y.B., M.D., H.A., F.Q., X.Y., X.L., M.A., X.Z., J.Z., X.C., C.Y., C.C., M.N., P.N., D.C., M.W., C.A. and F.X.

